# TSPO PET binding *in vivo* reflects increased phagocytic microglia at *post mortem* in people with frontotemporal dementia

**DOI:** 10.64898/2026.06.22.26356176

**Authors:** Davi S. Vontobel, Kei Onn Lai, Mehtap Bacioglu, George Nolan, Daniel C Maddison, Amelie Adamski, Julia Goddard, Noah L. Shapiro, Harry Crook, Tim Fryer, Young Hong, Sasvi Wijesinghe, Franklin Aigbirhio, Edward Avezov, Kieren S. J. Allinson, Annelies Quaegebeur, John T O’Brien, James B Rowe, Maria Grazia Spillantini, Maura Malpetti

## Abstract

Brain inflammation is a key feature of frontotemporal dementia (FTD). TSPO PET is widely used as an *in vivo* proxy for neuroinflammation, but whether the elevated signal reflects microglial, astrocytic, or vascular pathology is controversial. We paired *ante mortem* [¹¹C]PK11195 TSPO PET with *post mortem* neuropathology in 10 individuals with FTD (5 FTLD-tau, 5 FTLD-TDP) and 5 controls, combining CD68 immunohistochemistry across 17 regions, multiplex immunofluorescence pairing TSPO with microglial/macrophagic (IBA1, CD68), astrocytic (GFAP) and endothelial (CD31) markers, and three-dimensional single-cell reconstruction. CD68 burden was elevated in FTD, and correlated with regional TSPO PET binding across pathologies (β = 8.40, *P* < 0.001). The CD68-TSPO co-localised fraction tracked the PET signal, with CD68+ IBA1+ cells having increased TSPO expression over CD68-cells. The elevated TSPO PET signal in FTD likely reflects an increased burden of CD68+ microglia, supporting TSPO PET as a microglial-burden biomarker in both FTLD-tau and FTLD-TDP.

## Introduction

Frontotemporal dementia (FTD) encompasses a heterogeneous group of clinical syndromes, including behavioural variant FTD (bvFTD), semantic-variant primary progressive aphasia (svPPA), and non-fluent-variant primary progressive aphasia (nfvPPA) (Rascovsky et al., 2011; Gorno-Tempini et al., 2011). Within this clinical heterogeneity, there is limited correspondence between phenotype and underlying neuropathological subtypes of frontotemporal lobar degeneration (FTLD)(Grossman et al., 2023). The most common pathological substrates are primary tauopathies (3R or 4R tau, FTLD-tau) and TAR DNA-binding protein 43 (FTLD-TDP) (Neumann et al., 2021), with rarer cases due to fused-in-sarcoma (FUS) proteinopathy.

Brain inflammation has emerged as a central feature of many neurodegenerative diseases, including FTD and other syndromes associated with FTLD (Bright et al., 2019; Alster & Madetko-Alster 2025; Ishizawa & Dickson 2001). PET ligands targeting the 18 kDa translocator protein (TSPO) are the most widely used *in vivo* PET markers to index brain inflammation in patients with these conditions (Malpetti et al., 2024). A meta-analysis of 156 case-control studies confirmed elevated TSPO signal in core pathological regions across neurodegenerative conditions (De Picker et al., 2023). In sporadic and genetic FTD, there are disease-specific regional patterns of elevated TSPO PET binding (Malpetti et al., 2023; Malpetti et al., 2021). The three clinical subtypes differ in their neuroanatomical TSPO PET signal distribution (Bevan-Jones et al., 2020), while increased signal in frontal regions predicts faster cognitive decline across all variants (Malpetti et al., 2023). Elevated TSPO binding has also been observed across the broader FTLD spectrum (Kim et al., 2019), including the primary tauopathies progressive supranuclear palsy (PSP) and corticobasal degeneration (Palleis et al., 2021; Passamonti et al., 2018). Increased TSPO binding in disease-specific regions predicts clinical decline, and correlates with tau pathology and its longitudinal progression (Malpetti et al., 2020; Malpetti et al., 2021; Shapiro et al., 2025).

However, the cellular basis of TSPO brain PET signal elevation in neurodegenerative diseases remains poorly resolved. Within the central nervous system, TSPO is primarily expressed by microglia, astrocytes, and endothelial cells (Cosenza-Nashat et al., 2009; Guilarte et al., 2022), with rare reports of neuronal expression (Nutma et al., 2021). An established paradigm held that microglial populations overexpress TSPO in neuroinflammatory states (Guilarte et al., 2022; Fairley et al., 2024). Nutma et al. (2023) challenged this view using human post-mortem tissue from multiple neurodegenerative and neuroinflammatory diseases, including Alzheimer’s disease (AD), amyotrophic lateral sclerosis, and multiple sclerosis, and found no evidence of increased per-cell TSPO expression in microglia or astroglia in disease when using single immunohistochemical markers (ei, IBA1, CD68, HLA-DR). They concluded that the *in vivo* TSPO signal reflects changes in inflammatory cell density rather than per-cell upregulation. Work by Garland et al. (2024) in AD demonstrated that TSPO-expressing microglia are predominantly CD68+, linking the signal to a phagocytic microglial phenotype. In PSP, we recently reported a significant positive correlation between regional ^11^C-PK11195 binding and the increased burden of post-mortem CD68+ phagocytic microglia, and microglial TSPO levels, but not pathological astrocytic or endothelial TSPO (Wijesinghe et al., 2025).

The present study builds on this approach, to determine the cellular basis of TSPO PET signal elevation in FTD, directly linking *in vivo* PET binding with *post mortem* immunohistochemistry. In contrast to the clinically consistent PSP cohort with high clinicopathological correlation, we examine a clinically and pathologically heterogeneous FTD cohort with FTLD-tau and FTLD-TDP. We test the hypotheses that: (I) *in vivo* [¹¹C]PK11195 PET binding is associated with *post mortem* phagocytic microglia (specifically, CD68+) burden regardless of the underlying FTLD pathology; (II) *in vivo* TSPO PET signal reflects TSPO expression in phagocytic microglia over and above other cell types. The clinicopathological diversity of the cohort enables us to determine the pathological-specificity of the TSPO-microglial relationship.

## Results

### §1. Cohort characteristics

Demographic, clinical and neuropathological details for the 15 participants in this study are summarized in **Table 1**. Ten patients with a clinical diagnosis of FTD (3 females, 7 males; mean age at death 71.1 years, with bvFTD, svPPA and nfvPPA phenotypes) and five age- and sex-matched controls (2 females, 3 males; mean age at death 73.6 years) were included for post-mortem group comparisons. The two groups did not differ in age at death (Wilcoxon rank-sum test: W = 22, *P* = 0.713; Hodges-Lehmann shift FTD - Control = −2.0 years [95% CI −13.0, +10.0]) or sex distribution (Fisher’s exact test: *P* = 1.00). All ten patients underwent TSPO [¹¹C]PK11195 PET scans during their lifetime and subsequently donated their brains to the Cambridge Brain Bank; the mean ante-mortem PET-to-post-mortem interval was 66.7 ± 34.0 months (range 7 to 119 months). The death-to-formalin post-mortem interval was 64.1 ± 12.4 h in individuals with FTD (median 64, range 39-83) and 57.4 ± 22.3 h in controls (median 51, range 32-92); the groups did not significantly differ (Wilcoxon W = 33.5, *P* = 0.327).

**Table 1.** Demographic, clinical and neuropathological information for patients and controls. Patients with FTD listed first (n = 10, ordered by ascending age at scan), age- and sex-matched controls below (n = 5, ordered by ascending age at death). PET-PM = interval between TSPO PET scan and brain donation, in months.

| ID | Group | Sex | PET-PM (months) | Clinical syndrome | Post-mortem findings |
| --- | --- | --- | --- | --- | --- |
| Patient 1 | FTD | F | 91 | bvFTD | FTLD-tau (MAPT 10+16 mutation carrier) |
| Patient 2 | FTD | M | 124 | nfvPPA | FTLD-tau (CBD) |
| Patient 3 | FTD | M | 50 | bvFTD | FTLD-TDP Type B (C9orf72 mutation carrier) |
| Patient 4 | FTD | M | 56 | bvFTD | FTLD-tau (Pick's disease) |
| Patient 5 | FTD | F | 100 | svPPA | FTLD-TDP Type C |
| Patient 6 | FTD | M | 63 | nfvPPA | FTLD-TDP Type A (GRN mutation carrier) |
| Patient 7 | FTD | M | 54 | svPPA | FTLD-TDP Type C |
| Patient 8 | FTD | M | 86 | svPPA | FTLD-TDP Type C |
| Patient 9 | FTD | M | 36 | nfvPPA | FTLD-tau (PSP, Kovacs stage 6) |
| Patient 10 | FTD | F | 7 | nfvPPA | FTLD-tau (CBD) |
| Control 1 | Control | M | - | - | Normal brain (Braak I, CERAD 0, Thal phase 0) |
| Control 2 | Control | F | - | - | No significant neurodegenerative disease (Braak I; mild age-related amyloid angiopathy) |
| Control 3 | Control | M | - | - | No significant neuropathological changes |
| Control 4 | Control | F | - | - | No neurodegenerative disease (Braak II) |
| Control 5 | Control | M | - | - | No significant neurodegenerative pathology; Braak III tau, mild arterial sclerosis |

Neuropathological assessment confirmed substantial within-cohort variability: five patients carried FTLD-tau pathology and five carried FTLD-TDP pathology. Three of the ten patients were carriers of FTLD-related gene mutations: Patient 1 carried a *MAPT* mutation (c.915+16C>T, also coded as *MAPT* 10+16); Patient 3 carried a non-specific *C9orf72* mutation detected via TP-PCR with over 30 hexanucleotide repeats; and Patient 6 carried a c.388_391delCAGT, p.(Gln130Ser)fs *GRN* mutation. Controls were defined by the absence of significant neurodegenerative pathology at autopsy and Braak tangle stages ≤ III. Participant information provided in Table 1.

### §2. Phagocytic microglia burden (CD68+) is elevated in FTD

We tested whether CD68 burden at *post mortem* differed between patients with FTD and controls using a linear mixed-effects model (LMEM) with group (FTD vs controls), region, and their interaction as fixed effects, and a random intercept for subject; *CD68 ∼ Group × Region + (1|id)*. Type III F-tests with Kenward-Roger degrees of freedom were applied across 16 cortical, subcortical, brainstem and cerebellar regions (BA4 excluded due to lack of control coverage). CD68 area fraction was elevated in patients with FTD compared to controls (group main effect: F(1, 13.9) = 14.63, *P* = 0.002; β_(FTD-Ctrl) = +2.21, marginal R² = 0.22; Fig. 1).

**Figure 1.**
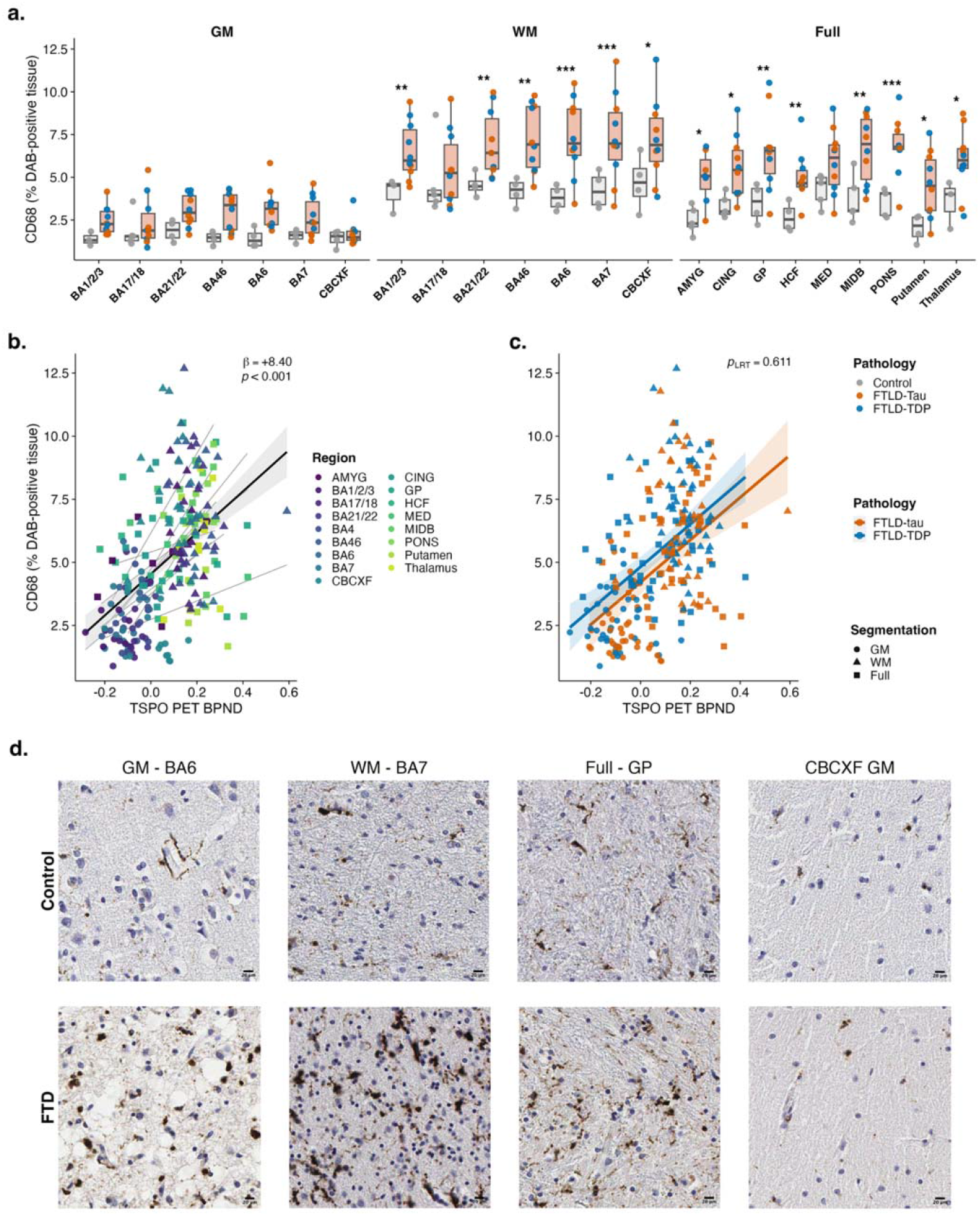
Post-mortem CD68 burden by anatomical region in patients with FTD vs controls, and its coupling with regional *in vivo* TSPO PET binding. (A) CD68 %DAB tissue area in patients with FTD vs Controls, (grey matter and white matter facets: BA1/2/3, BA17/18, BA21/22, BA46, BA6, BA7, CBCXF = cerebellum; Full = non-segmented facet: AMYG = amygdala, CING = cingulate, GP = globus pallidus, HCF = hippocampal cortex, MED = medulla, MIDB = midbrain, Pons, Putamen, Thalamus). Asterisks mark FTD-vs-Control contrasts at *P* <0.05 *, *P* <0.01 **, *P* <0.001 ***. BA4 was excluded from the Control x FTD comparison due to absent control coverage; CD68 ∼ Group × Region + (1|id), Type III F-tests with Kenward–Roger df: group main effect F(1, 13.9) = 14.63, *P* = 0.002, β_(FTD−Ctrl) = +2.21, marginal R² = 0.22; Group × Region F(15, 279.1) = 0.47, *P* = 0.956; N = 323 observations / 15 subjects. (B) Regional CD68 area fraction vs regional TSPO PET binding (BPND) in all 10 patients with FTD; per-subject linear fits (grey) overlaid with the overall LMEM trend (black, 95% CI ribbon); CD68 ∼ BPND + PET_PM_Interval + (1|id): β_BPND = +8.40, SE = 0.84, t = 9.96, *P* < 0.001; marginal R² = 0.31, conditional R² = 0.45; N = 247 observations / 10 subjects. (C) Same LMEM as panel B with pathology-specific fits overlaid; CD68 ∼ PathClass × BPND + PET_PM_Interval + (1|id): slope-equality LRT χ² = 0.258, df = 1, *P* = 0.611; FTLD-tau β = 8.91, FTLD-TDP β = 7.97. (D) Representative CD68 DAB-immunohistochemistry images, one FTD case and one matched control per region; scale bar 20 μm. From left to right: grey-matter BA6, white-matter BA7, globus pallidus, and cerebellar grey matter, scale bar = 20 μm.

Post hoc analyses with a segmentation-aware companion model (*CD68 ∼ Group × Region × Segmentation + (1|id)* identified regions where CD68 area fraction was higher in patients than controls (emmeans package; see Table 2 for per-region contrasts). Across cortical Brodmann regions, elevation in CD68 area fraction in patients was predominantly in white-matter: CD68 differences in white-matter were significant in BA6, BA7, BA21/22, BA46, BA1/2/3 and the cerebellum (CBCXF), while in contrast no cortical grey matter contrast reached significance (*P* > 0.05). Concordantly, CD68 area fraction in cerebellum was higher in patients than controls in white matter regions (*P* = 0.011) but not in grey matter (*P* = 0.808). Primary visual cortex (BA17/18) did not show a consistent group difference in grey-matter or white-matter areas. When analyzing the full-quantified regions (no segmentation between white and grey matter), we found increased CD68 burden in the anterior cingulate, globus pallidus, hippocampal cortex, putamen, thalamus, amygdala, midbrain and pons, with medulla at trend (*P* = 0.065).

**Table 2.**
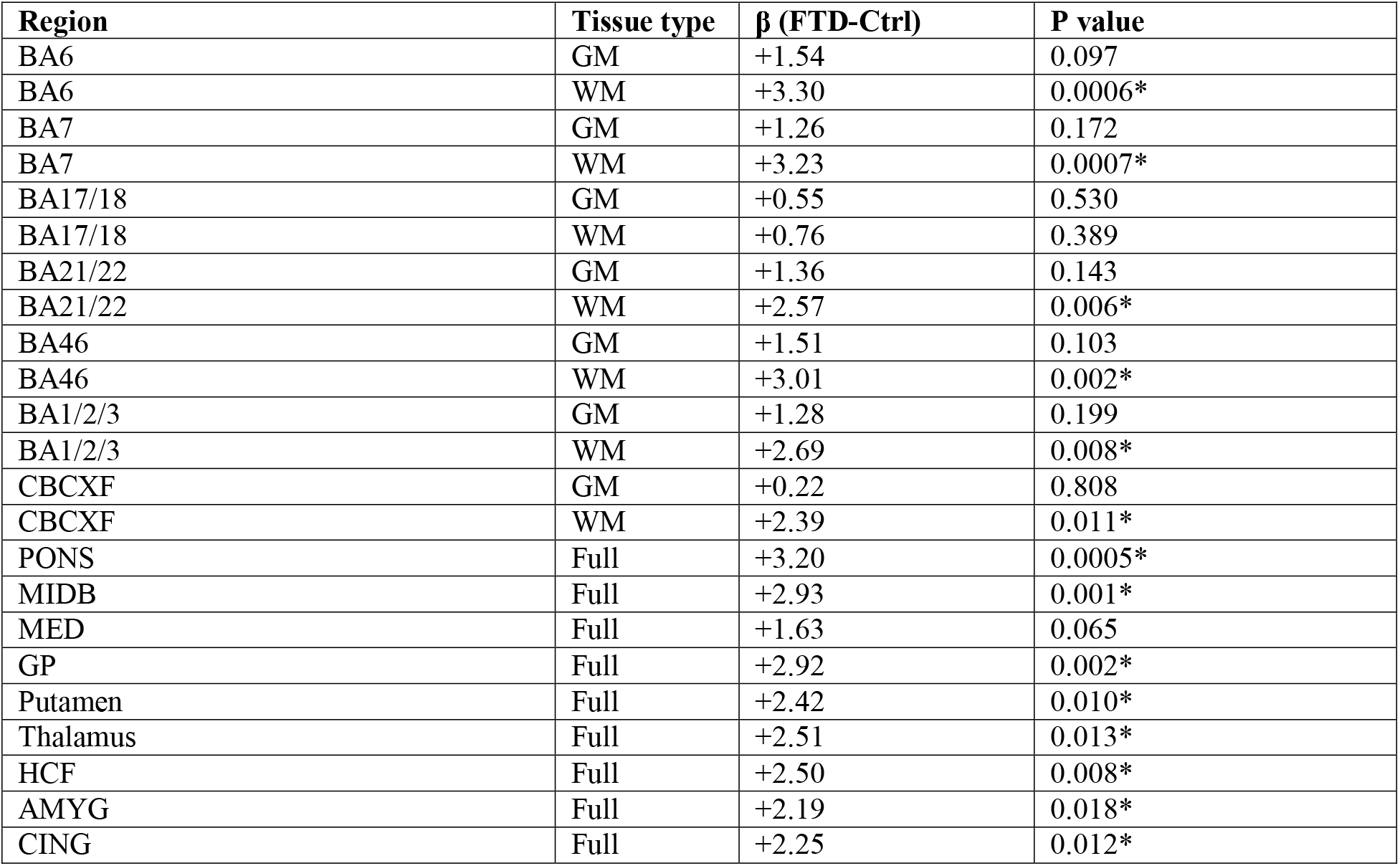
Group comparisons on CD68 area fraction values: results from linear mixed-effects model (Region × Segmentation x Groups). Significance grades reflect linear mixed-effects model posthoc emmeans-contrasts: * *P* < 0.05. Tissue type: GM = grey matter, WM = white matter, Full = non-segmented sampling.

### §3. *In vivo* TSPO PET binding correlates with *post mortem* phagocytic microglial markers

To determine whether TSPO PET binding potential (BPND) reflects phagocytic microglial/macrophage burden (CD68+ at post-mortem), a LMEM modelled CD68 area fraction as the outcome with regional BPND and PET-to-post-mortem interval as fixed-effect predictors and a subject-level random intercept (*CD68 Area Fraction ∼ TSPO PET BPND + PET_PM_Interval + (1|id)*). Across all 10 patients with FTD, the TSPO PET signals were positively associated with CD68 area fraction (β = 8.40, SE = 0.84, t = 9.96, *P* < 0.001; marginal R² = 0.31, conditional R² = 0.45; Fig. 1B). The PET-to-post-mortem interval was not a significant predictor.

To test whether the *ante mortem* TSPO-signal-CD68 association differed by pathology, we extended the model with an interaction between regional BPND and pathology subgroup (FTLD-tau vs FTLD-TDP; *CD68 ∼ PathClass × BPND + PET_PM_Interval + (1|id)*; slope-equality LRT against the no-interaction reduced model *CD68 ∼ PathClass + BPND + PET_PM_Interval + (1|id)).* The interaction did not improve fit (LRT χ² = 0.258, df = 1, *P* = 0.611), and per-pathology slopes were comparable for FTLD-tau and FTLD-TDP cases (FTLD-tau β = 8.91, 95% CI [6.53, 11.29]; FTLD-TDP β = 7.97, 95% CI [5.64, 10.31]; slope contrast TDP - Tau = −0.94, *P* = 0.581; Fig. 1C). The TSPO PET-CD68 coupling is therefore observed across both proteinopathies with comparable magnitude. As a follow up analysis we reused neuropathological diagnosis slides (AT8 pTau and pTDP-43 stains) from the cohort for verification with the TSPO PET signal and CD68 staining. TSPO PET did not meaningfully correlate with protein aggregate load in either FTLD subgroup; however, AT8+ tau and CD68 displayed a mild positive association pre FDR correction (Fig. S1A).

### §4. Tissue TSPO area is elevated in FTD and correlates with regional TSPO PET binding

Given the positive association between *in vivo* TPSO PET and *post mortem* CD68, we tested whether total tissue TSPO area at *post mortem* was elevated in people with FTD relative to controls and associated with regional TSPO PET *in vivo* signal. Two vicinal slide sets per subject (Set 1: IBA1/CD68/TSPO; Set 2: GFAP/CD31/TSPO) were quantified across BA6, BA21/22 and BA46, with TSPO area expressed as ratio of TSPO+ area over total tissue area (TSPO area fraction = TSPO area / Total area) and analyzed independently per slide set. Model formula: per-slide-set FTD-vs-Control *Norm_AreaB ∼ Group + Region + (1|id)*.

TSPO area fraction was elevated in patients with FTD relative to controls in both slide sets (Set 1, IBA1/CD68: β_(FTD-Ctrl) = +0.013, *P* = 0.014, *P*_FDR = 0.014; Set 2, GFAP/CD31: β = +0.022, *P* = 0.008, *P*_FDR = 0.014; Fig. 2A-B). The two slide sets agreed in direction and magnitude despite being processed independently. The Region main effect was not significant in either slide set (Set 1, IBA1/CD68: F(2, 26.4) = 1.11, *P* = 0.344; Set 2, GFAP/CD31: F(2, 26.1) = 1.31, *P* = 0.287).

**Figure 2.**
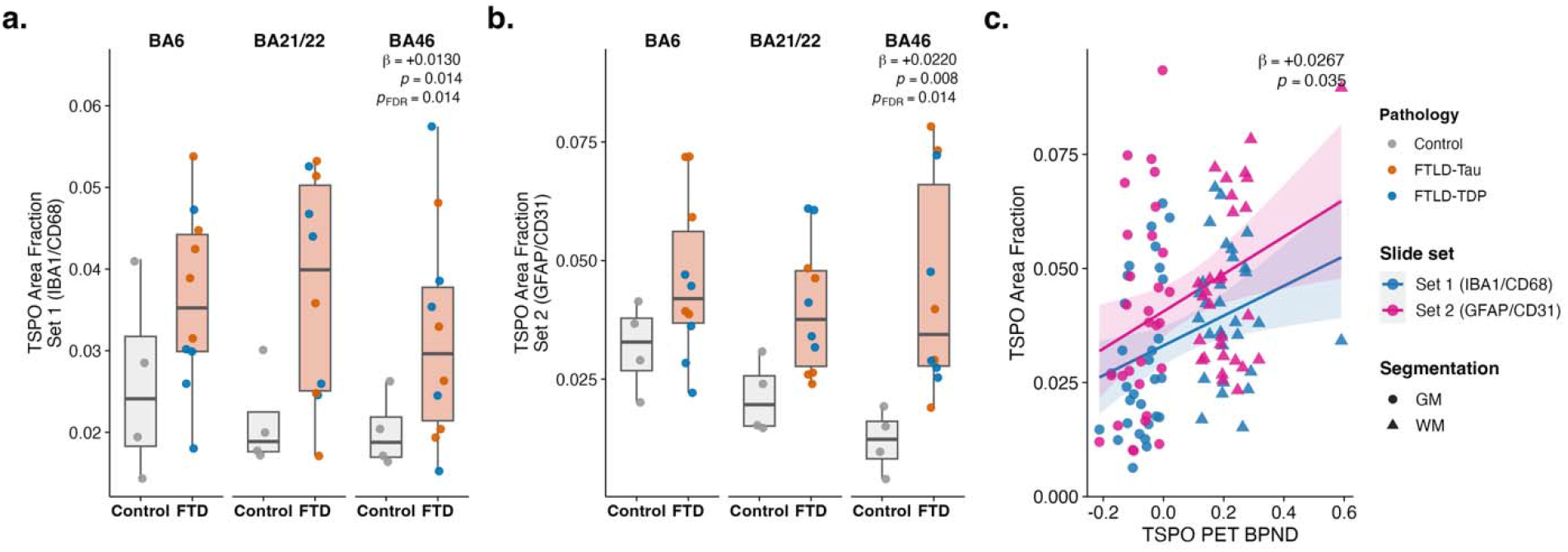
Post-mortem tissue TSPO area fraction is increased in frontotemporal regions and tracks regional *in vivo* TSPO PET binding in FTD. (A) FTD-vs-Control contrast for Set 1 (IBA1/CD68 stain); box fill salmon = FTD, light grey = Control; points coloured by pathology subgroup; Set 1 (IBA1/CD68): β = +0.0130, *P* = 0.014, *P*_FDR = 0.014. (B) FTD-vs-Control contrast for Set 2 (GFAP/CD31 stain); same encoding and model as panel A; Set 2 (GFAP/CD31): β = +0.0220, *P* = 0.008, *P*_FDR = 0.014. (C) Joint paired-subject TSPO-area × TSPO PET scatter across all 10 patients with FTD, both slide sets overlaid on the same axes; ribbon colour = slide set (Set 1 IBA1/CD68 = blue, Set 2 GFAP/CD31 = magenta); shape = segmentation (grey matter = circle, white matter = triangle); Norm_AreaB ∼ BPND × SlideSet + PET_PM_Interval + (1|id); Set 1 β = +0.0267, *P* = 0.036, *P*_FDR = 0.036. Set 2 (GFAP/CD31) β = +0.0348, *P* = 0.007, *P*_FDR = 0.013.

Within FTD, *post mortem* regional TSPO area fraction was positively associated with regional TSPO PET signal in both slide sets, jointly modelled joint paired-subject BPND coupling *Norm_AreaB ∼* TSPO PET *× SlideSet + PET_PM_Interval + (1|id)* (Set 1, IBA1/CD68: β_BPND = +0.0267, *P* = 0.036, *P*_FDR = 0.036; Set 2, GFAP/CD31: β_BPND = +0.0348, *P* = 0.007, *P*_FDR = 0.013; slope contrast Set 1 - Set 2 = −0.008, *P* = 0.642; Fig. 2C). PET-to-post-mortem interval was a covariate of no interest.

### §5. TSPO expression co-localises with phagocytic microglia

We then tested whether co-localisation of immunofluorescence signals between TSPO and each of four cell-type-specific markers are elevated in the FTD group. These markers include IBA1 (microglia/macrophage membrane), CD68 (microglia/macrophages lysosomes), GFAP (astroglial cytoskeleton), and CD31 (vascular endothelial cells/blood vessels). Image-level co-localised area (TSPO and marker double-positive area / total tissue area; ratio of co-localised area over total area) was modelled per marker by a binary LMEM (FTD vs Control) with region as a fixed effect and a subject random intercept (*Coloc/Total ∼ Group + Region + (1|id)*).

Area fraction of co-localisation with TSPO was elevated in patients with FTD relative to controls for three markers (CD68: β_(FTD-Ctrl) = +0.0012, *P* = 0.024, *P*_FDR = 0.048; IBA1: β = +0.0038, *P* = 0.022, *P*_FDR = 0.048; GFAP: β = +0.0036, *P* = 0.042, *P*_FDR = 0.056; Fig. 3 C– F), while the endothelial CD31-TSPO co-localised area was not (CD31: β = +0.0035, *P* = 0.071, *P*_FDR = 0.071). However, with multiple comparison correction across the 4-marker family, comparisons in TSPO co-localisation with CD68 and IBA1 markers remained significant (*P*_FDR = 0.048 each), while TSPO co-localisation areas with GFAP and CD31 markers were not statistically significant (p>0.05).

**Figure 3.**
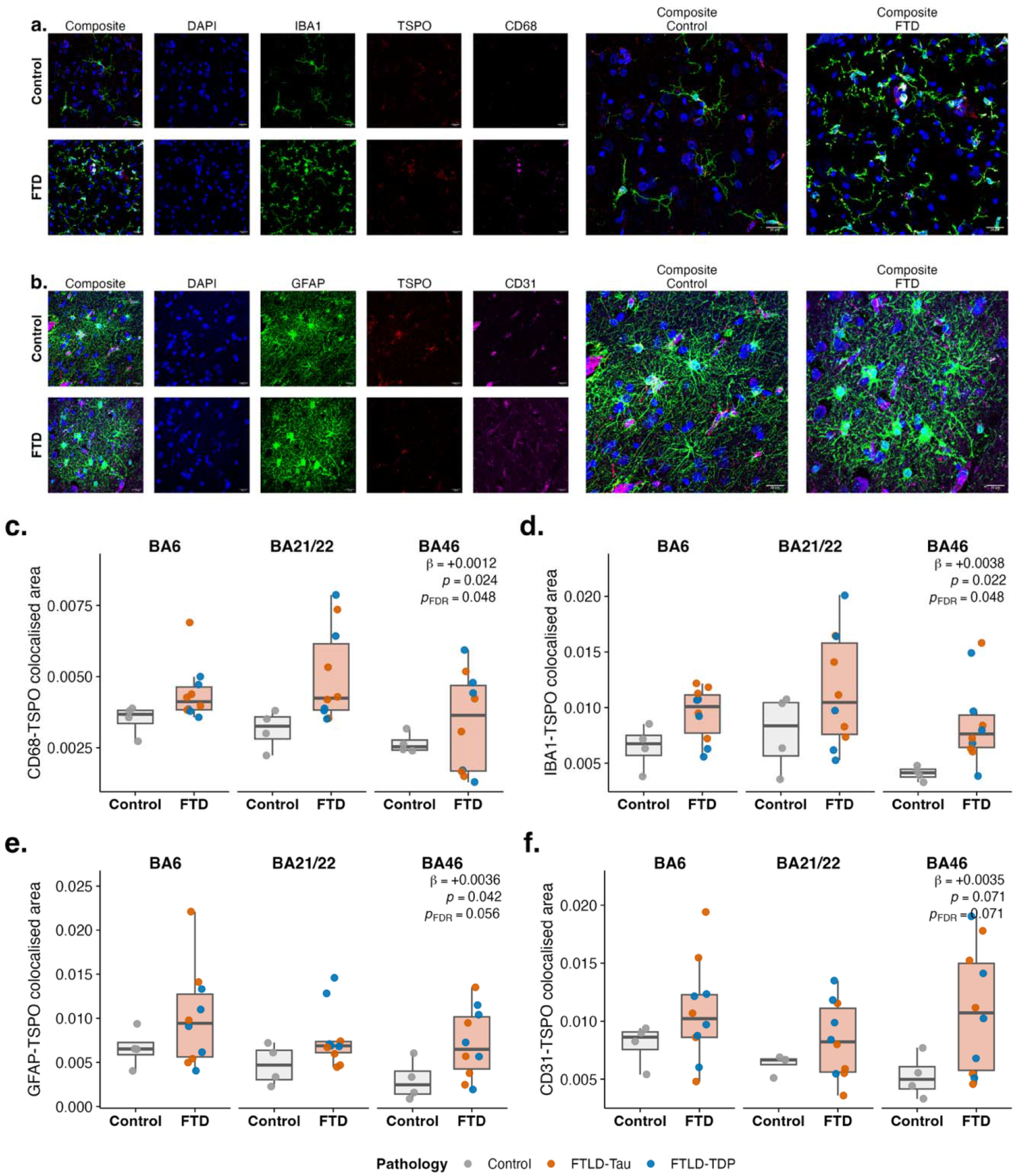
Marker-TSPO co-localised area is elevated in FTD in microglia. (A) Representative immunofluorescence in BA46 in FTD and matched controls across DAPI / IBA1 / TSPO / CD68 single-channel columns and the merged composite; scale bar = 20 μm. (B) Representative immunofluorescence in BA46, same layout as panel A for DAPI / GFAP / TSPO / CD31; scale bar = 20 μm. (C–F) Coloc_by_Total ∼ FTD_vs_Control + Region + (1|id); FTD−Control β per marker with BH-FDR across the 4-marker family. (C) CD68-TSPO co-localised area (Coloc / Total Area) compared between FTD and Controls in BA6, BA21/22 and BA46; box fill salmon = FTD, light grey = Control; points coloured by pathology subgroup; CD68-TSPO: β = +0.0012, *P* = 0.024, *P*_FDR = 0.048. (D) IBA1-TSPO co-localised area; IBA1-TSPO: β = +0.0038, *P* = 0.022, *P*_FDR = 0.048. (E) GFAP-TSPO co-localised area; GFAP-TSPO: β = +0.0036, *P* = 0.042, *P*_FDR = 0.056. (F) CD31-TSPO co-localised area; CD31-TSPO: β = +0.0035, *P* = 0.071, *P*_FDR = 0.071.

### §6. TSPO PET binding only correlated with phagocytic microglia burden

To identify which cellular sources of TSPO drive the *in vivo* PET signal in patients with FTD, we tested the association between co-localisation area for each markers and regional TSPO PET signal, fitting a LMEM per marker across BA6, BA21/22 and BA46. The random-effects structure was selected by the Matuschek ladder per marker. Model formula: *Co-localised Area Fraction ∼ TSPO PET BPND + PET_PM_Interval + Random Effects*.

Across all patients, regional TSPO PET signal was positively associated with phagocytic microglia (CD68-TSPO co-localised fraction: β = +0.0041, *P* = 0.015, *P*_FDR = 0.062) and negatively associated with the endothelial CD31-TSPO co-localised fraction (β = −0.0073, *P* = 0.031, *P*_FDR = 0.062) before FDR correction, with a graphical-trend-level negative association for IBA1-TSPO (β = −0.0072, *P* = 0.084, *P*_FDR = 0.112) and no association for GFAP-TSPO (β = −0.0008, *P* = 0.90, *P*_FDR = 0.90; Fig. 4).

**Figure 4.**
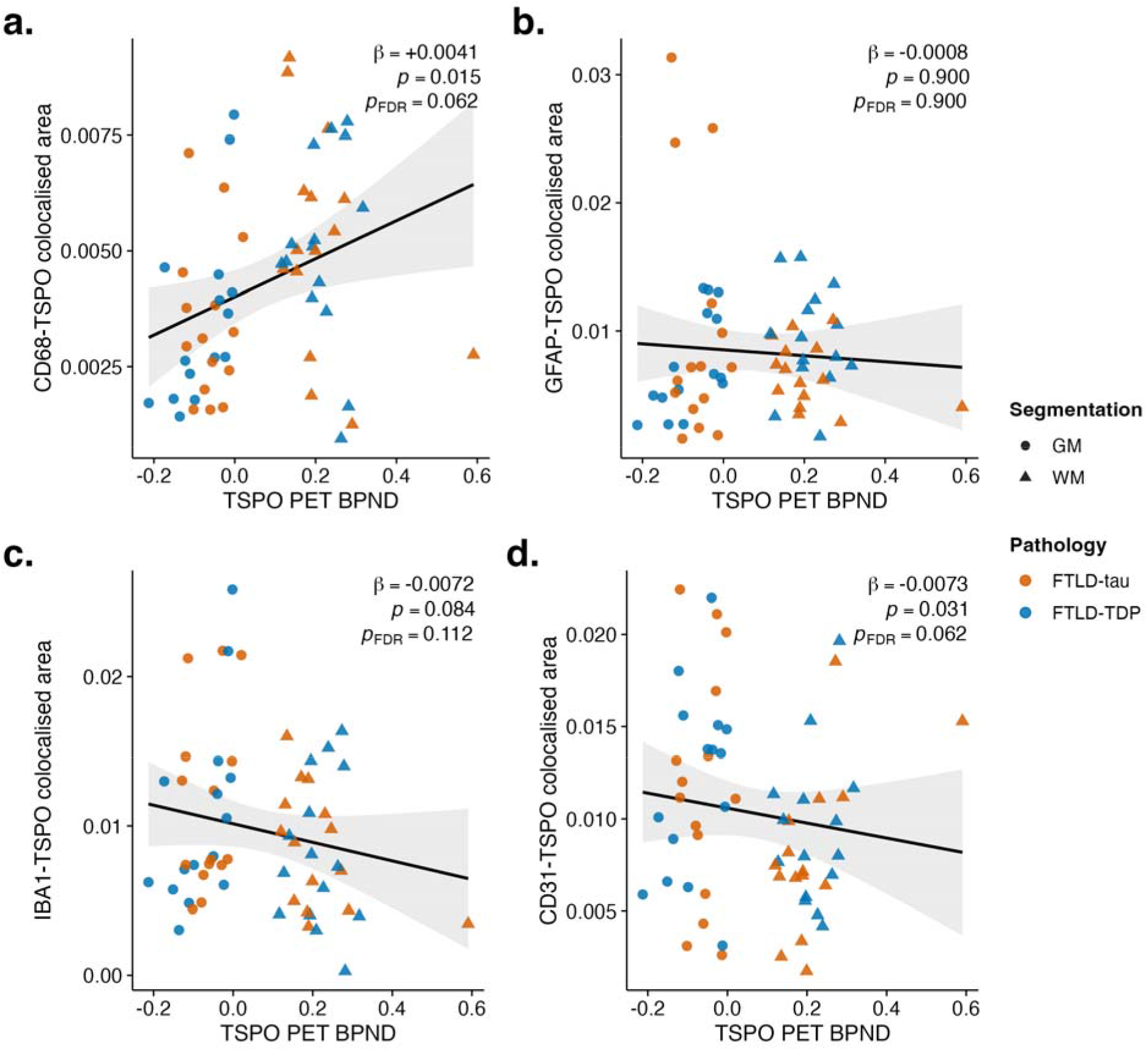
Cell-type-specific marker-TSPO co-localised area modelled against regional TSPO PET in FTD. Coloc / Total Area vs regional BPND in all 10 patients with FTD, fitted per marker. Points coloured by pathology subgroup (FTLD-tau orange, FTLD-TDP blue); shape = segmentation (grey matter = circle, white matter = triangle, Full = square), (A–D) Coloc_by_Total ∼ BPND + PET_PM_Interval + random effects, fitted per marker with Matuschek-selected structure (CD68, IBA1, CD31 = (1|id); GFAP = (1 + BPND|id)); BH-FDR across the 4-marker family.. (A) CD68-TSPO co-localisation; β_BPND = +0.0041, *P* = 0.015, *P*_FDR = 0.062. (B) GFAP-TSPO co-localisation; β_BPND = −0.0008, *P* = 0.900, *P*_FDR = 0.900. (C) IBA1-TSPO co-localisation; β_BPND = −0.0072, *P* = 0.084, *P*_FDR = 0.112. (D) CD31-TSPO co-localisation; β_BPND = −0.0073, *P* = 0.031, *P*_FDR = 0.062.

The four-marker pattern positions phagocytic microglia (CD68+) as the cell type marker whose co-localisation with TSPO tracks the *in vivo* PET signal positively, with null or negative associations for IBA1, GFAP, and CD31. A grey-matter / white-matter decomposition (Supplementary Figure S3) shows the CD68 positive coupling concentrated in grey matter (GM β = +0.013, *P* = 0.052; WM β = −0.009, *P* = 0.027; TSPO PET signal × Segmentation interaction LRT *P* = 0.007), with the same compartment-specific structure for IBA1 (GM β = +0.048, *P* = 0.006; WM β = −0.016, *P* = 0.122; LRT *P* = 0.001). GFAP and CD31 show no significant Segmentation interaction (LRT *P* = 0.310 and 0.88 respectively).

### §7. Single-cell metrics of microglial TSPO in FTD

To quantify TSPO expression per microglial cell, we performed 3D rendering of IBA1+ cells in confocal images. Variables were aggregated to per-subject means across quantification samples. Total TSPO volume per cell was acquired. Within IBA1+ microglia we classified each cell as CD68− or CD68+ (CD68 volume 0 vs > 0) and compared, per subject, the mean TSPO per cell, the TSPO fraction of cell volume, and cell size, analyzing Control and FTD separately (Fig. 5B– G).

**Figure 5.**
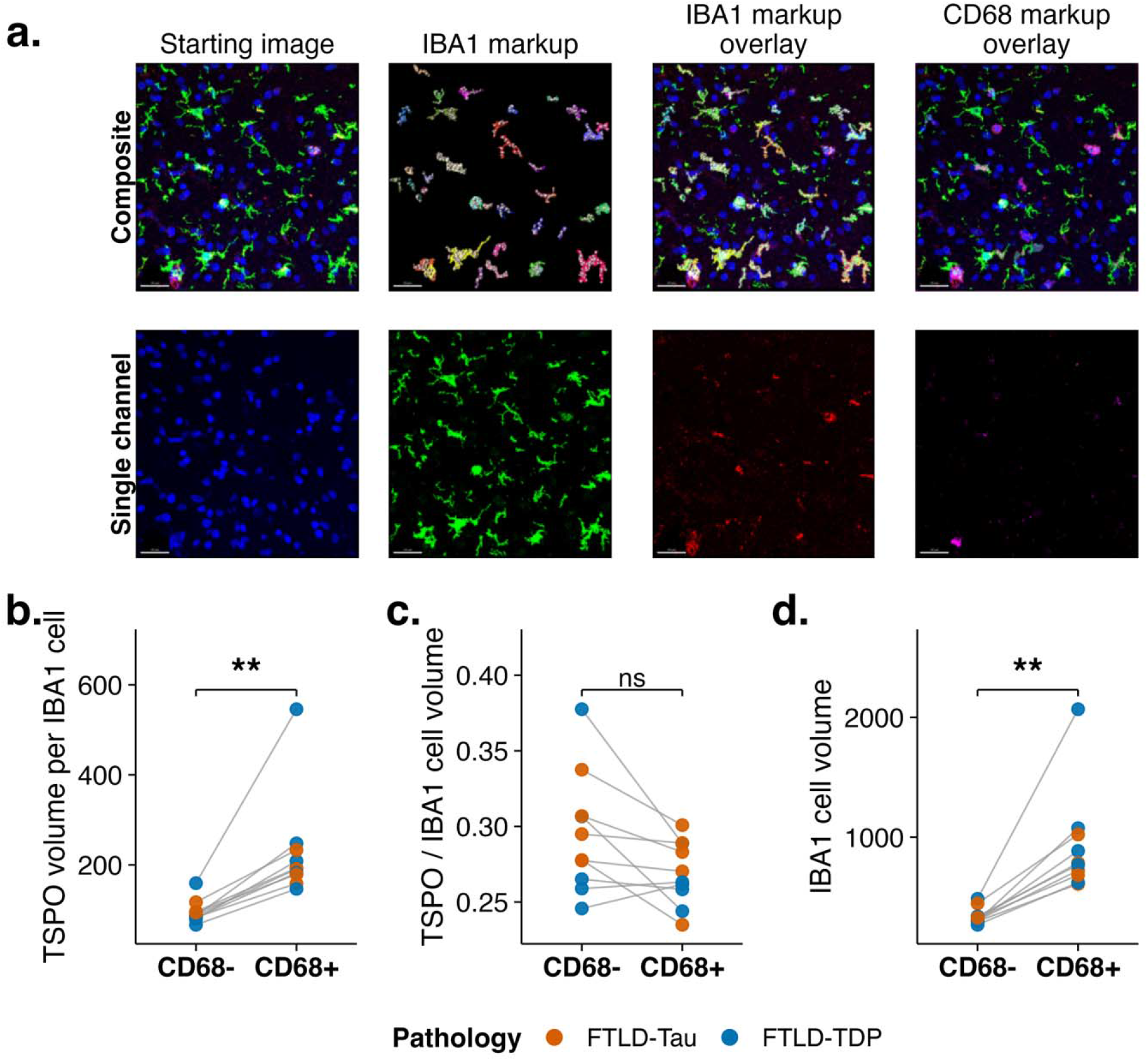
CD68+ microglia carry more TSPO in proportion to their size. Colour palette throughout: Control = grey, FTLD-tau = orange, FTLD-TDP = blue. (A) Representative Imaris three-dimensional single-cell segmentation from BA46 tissue from an FTD case: an IBA1+ cell-body surface with intracellular TSPO and CD68 vesicle objects rendered within the cell volume; scale bar = 30 μm. (B–D) Within IBA1+ microglia, each cell was classified CD68− or CD68+ (CD68 volume 0 vs > 0) and per-subject means compared between classes by paired Wilcoxon signed-rank test, analysing Control (B–D, n = 4) and FTD (E–G, n = 10) separately. (B) mean TSPO volume per IBA1+ cell; (C) TSPO fraction of cell volume; (D) IBA1+ cell volume. Each grey line links one subject’s CD68− and CD68+ means; points coloured by pathology subgroup. Brackets show BH-FDR–adjusted significance within each metric (** P_FDR < 0.01; ns, not significant). In FTD, CD68+ microglia carried more TSPO per cell (10/10 subjects; V = 55, P = 0.002, P_FDR = 0.004) and were larger (10/10; V = 55, P = 0.002, P_FDR = 0.004), the TSPO fraction of cell volume was not higher (8/10; V = 6, P = 0.027, P_FDR = 0.055, ns).

In FTD, CD68+ microglia carried more absolute TSPO than CD68− microglia (10/10 subjects; *P* = 0.002, *P*_FDR = 0.004) and were larger (10/10; *P* = 0.002, *P*_FDR = 0.004), whereas the TSPO fraction of cell volume was not higher — if anything lower (8/10; *V* = 6, *P* = 0.027, *P*_FDR = 0.055, ns). Controls were concordant in direction for every metric but did not reach significance at n = 4 (*P > 0.05*). IBA1+ cell counts in the sampled fields were equivalent between groups (*P* = 0.288; Figure S5), excluding a sampling or cell-density bias within the fields.

## Discussion

The principal finding of this study is that in both FTLD-tau and FTLD-TDP forms of frontotemporal dementia, increased TSPO PET binding is associated with CD68+ phagocytic microglia. *Post mortem* TSPO signal was also tightly linked with the *in vivo* TSPO PET signal, across patients and brain regions. Overall, there was an increase in microglial area (both IBA1+ and CD68+) co-localised with TSPO, while astrocytic and endothelial TSPO remained comparable to controls (arguable trend in GFAP+ astrocytes). However, only the CD68-TSPO area was positively linked with the *in vivo* TSPO PET signal (before FDR correction). Finally, within BA46 exclusively, we analyzed the intracellular contents of the IBA1+ cells and found that CD68+ cells were both larger than their CD68-counterparts and expressed more absolute TSPO.

Elevated phagocytic-microglial protein burden at *post mortem* is a recurring neuropathological feature across neurodegenerative conditions. CD68-immunoreactive microglia are increased in Alzheimer’s disease (Hopperton et al., 2018), Parkinson’s disease (Doorn et al., 2014), and all major subtypes of frontotemporal lobar degeneration (Woollacott et al., 2020). Our finding of increased CD68 burden in both FTLD-tau and FTLD-TDP accords with this literature and extends it to a directly PET-matched FTD cohort at post-mortem. The positive association between *in vivo* TSPO PET signal and CD68 area fraction at *post mortem* across the brain expands the previous PET-to-*post mortem* evidence in PSP, where we reported strong positive relationship between TSPO PET and CD68 burden (Wijesinghe et al., 2025). In the present FTD cohort, the random-effects model selected by AIC similarly retained a subject-level intercept only (Matuschek et al., 2017). The preserved directionality and comparable magnitude of the association between *in vivo* TSPO PET BPND and post-mortem TSPO expression across the two independent cohorts suggests that the relationship may generalize across all FTLD subtypes, including 4R-tau PSP and in our heterogeneous FTLD-tau + FTLD-TDP. Previous *post mortem* analysis in AD demonstrated that the dominant TSPO sources within the temporal lobe were CD68+ microglia (Garland et al., 2024). These studies in combination suggest that the TSPO PET signal captures a pathology-independent microglial component across different neurodegenerative diseases.

The concordance between *ante mortem* TSPO PET binding and *post mortem* TSPO-immunoreactive area fraction positions TSPO as a marker of sustained neuroinflammatory burden rather than a transient signal that resolves with disease progression in FTD. Previous longitudinal TSPO PET studies have demonstrated stable signals over one-year intervals in multiple sclerosis white matter hyperintensities (Nylund et al., 2025) or increasing signals at follow up in AD in disease-associated areas (Fan et al., 2015; Kreisl et al., 2016). This contrasts with transient elevation following acute cerebrovascular injury (Jensen et al., 2025), illustrating important, if a dash obvious, differences between chronic brain inflammation in neurodegeneration and the acute inflammatory responses to discrete insults.

The fluorescent co-localisation analyses showed that the TSPO PET signal is preferentially associated with CD68+ microglia. Despite an observed increase in both CD68-TSPO and IBA1-TSPO areas within the FTD cohort, CD68-TSPO co-localisation was positively associated with regional TSPO PET *in vivo* signal, while GFAP-TSPO and IBA1-TSPO co-localised area fractions were not. Although this CD68–TSPO association did not survive multiple-comparison correction (P_FDR = 0.062), it recapitulated the larger 17-region CD68 burden association with in vivo TSPO PET signal. In this context, our findings suggest that the *in vivo* TSPO PET signal in patients with FTD is driven by phagocytic microglia, rather than TSPO expression in other cell types. This pattern is concordant with previous post-mortem-only observations in AD, where CD68-TSPO co-localisation was higher than the TSPO co-localisation with IBA1, HLA-DR, MSR-A, and CD64+ microglia/macrophages in the temporal cortex (Garland et al., 2024). The relatively weak negative association between TSPO PET binding and the CD31-TSPO co-localised area fraction may reflect age- and disease-related loss of endothelial TSPO binding sites (Tomasi et al., 2008; Rizzo et al., 2019); however, further quantitative pathology studies are needed to comprehend this signal decoupling.

Our data also highlight a clear dissociation between grey and white matter in the disease-level CD68 burden and the PET-coupling signal. The CD68 elevation in FTD cerebrum is white-matter driven: CD68 was significantly elevated in the white matter underlying every Brodmann region except primary visual cortex, as well as the cerebellum, whereas no cortical grey matter contrast reached significance. This pattern is consistent with the white matter neuroinflammatory burden increasingly recognized in FTLD and other neurodegenerative diseases (Woollacott et al., 2020). However, the association between TSPO PET and co-localised area was grey matter driven for both CD68 and IBA1. The TSPO PET x compartment interaction was significant for both markers, confirming the grey-vs-white matter dissociation. The tissue-wide IBA1 to TSPO PET divergence observed appears to be a white-matter-specific feature, consistent with reports that IBA1 immunoreactivity is sensitive to pathology (Hendrickx et al., 2017; Woollacott et al., 2020).

When interpreting these results, it is important to keep in mind that CD68 staining marks the lysosomal compartment in these cells (Quick et al., 2023). In this context, our results suggest that *in vivo* TSPO PET signal is enriched in association with a microglial/macrophage population with elevated CD68 signals, rather than a tissue-wide expansion of microglial density. Single-cell Imaris data support this interpretation: we found that IBA1+ cells which were also CD68+ were both larger in volume and expressed more absolute TSPO than CD68-cells, though the IBA1-TSPO volume ratio remained unchanged. This is consistent with recent studies which compared TSPO expression in single-marker labeled microglia: Nutma et al. (2023) showed that in human neurodegenerative tissue microglial TSPO was not up-regulated as a fraction of cell area when comparing cells labeled with IBA1, CD68 or HLA-DR. Our previous work on PSP reached the same conclusion, attributing the disease-related TSPO increase to microglial burden rather than per-cell up-regulation (Wijesinghe et al., 2025). However, in both studies, TSPO was only analyzed relative to the cell marker area. Here we report an increase of absolute TSPO in populations detected by IBA1 and CD68 in FTD suggesting an increased expression of TSPO in microglial cells with a phagocytic phenotype. Garland et al. (2024) also found CD68+ microglia to be the dominant TSPO-expressing population in Alzheimer’s disease, in keeping with our observation that CD68+ microglia are preferentially associated with TSPO. Set against the greater CD68 burden in the pathological groups, the concordance between this *post mortem* signal and *in vivo* TSPO PET, and the persistence of elevated TSPO to the point of death, these findings position CD68+ microglia as the cells most closely associated with the in vivo TSPO signal within our experiments.

As an exploratory analysis in pathology-specific sub-cohorts, we tested the association between phagocytic microglial burden (CD68) and tau or TDP-43 burden at post-mortem. Within the FTLD-tau sub-cohort, tau burden was modestly associated with CD68 burden across brain regions pre FDR correction. There was no association between pTDP-43 and CD68 in the FTLD-TDP sub-cohort. The CD68-tau coupling is partially concordant with prior *in vivo* TSPO-tau correspondences observed in multitracer PET studies in patients with primary tauopathies (Bevan-Jones et al., 2020; Malpetti et al., 2024; Appleton et al., 2025). In these studies, TSPO PET signal was regionally correlated with tau PET binding; however, the TSPO signals often extended beyond the tau epicenters. The absence of a CD68-pTDP-43 coupling does not discard a temporally contingent *in vivo* TSPO-TDP relationship, as TDP-43 PET tracers remain in development (Vokali et al., 2025). Of note, these exploratory analyses in pathology-specific sub-cohorts need to be interpreted with caution given the small per-subgroup sample sizes.

Our study has several limitations. Due to the rarity of post-mortem tissue availability in individuals with TSPO PET scans, while the sample sizes are larger than other PET-to-post-mortem studies this can still limit the power to detect small effects, and limits the interpretation of null findings. We accounted for this in several ways, including the implementation of linear mixed-effects models of all regional data points, and the use of FDR correction for interpretational robustness. [¹¹C]PK11195 is a first-generation TSPO tracer with a lower signal-to-noise ratio compared to second- and third-generation TSPO tracers (Malpetti et al., 2024). However, these alternative tracers are confounded by common genetic polymorphisms that substantially affect binding, making them less suitable for studies of rare diseases where gene-stratified recruitment would be especially challenging. Further studies could aim to replicate our approach with other TSPO tracers. The antibodies used carry their own constraints as IBA1 immunoreactivity purportedly degrades in disease-affected tissue (Hendrickx et al., 2017), and CD68 identifies the lysosome-enriched microglial compartment without completely resolving specific functional sub-states (Paolicelli et al., 2022), or inferences regarding lysosomal functional integrity (Quick et al., 2023). Expanding PET-to-post-mortem studies with novel TSPO tracers and deeper microglial phenotyping will be necessary to resolve the specific cellular programs captured by elevated TSPO PET signals in neurodegeneration.

In conclusion, we have shown that the elevated [¹¹C]PK11195 signal reflects increased CD68-immunoreactive and phagocytic microglial signal in both FTLD-tau and FTLD-TDP forms of frontotemporal dementia. TSPO+ microglial cells contribute to *in vivo* regional TSPO PET signal, over and above other cell types. We suggest that TSPO PET can be interpreted as a microglial signal in FTD and related disorders, empowering experimental medicine studies and clinical trials of disease-modifying therapies.

## Methods

### Participants

Written informed consent was obtained from all participants. The protocols were approved by the National Research Ethics Service’s East of England Cambridge Central Committee, and the UK Administration of Radioactive Substances Advisory Committee (Neuropathology Research in Dementia protocol, Research Ethics Committee reference 16/WA/0240; Neuroimaging of Inflammation in Memory and Related Other Disorders (NIMROD), Research Ethics Committee reference 13/EE/0104).

Ten people with clinical diagnosis of FTD were included (Table 1); all underwent [¹¹C]PK11195 PET scans during their lifetime and subsequently donated their brains to the Cambridge Brain Bank. Patients were clinically diagnosed with bvFTD (n = 3), nfvPPA (n = 4), or svPPA (n = 3) (Rascovsky et al., 2011; Gorno-Tempini et al., 2011). Neuropathological assessments revealed the expected variability within the cohort: 5 cases of FTLD-tau (4R tau: corticobasal degeneration n = 2, PSP n = 1; 3R tau: Pick’s disease n = 1; FTLD-tau-other n = 1) and 5 cases of FTLD-TDP (TDP-43 Type A n = 1, Type B n = 1, Type C n = 3). Three of the ten patients were carriers of FTD-related gene mutations: patient 1 carried a mutation in the *MAPT* gene (c.915+16C>T, also coded as the *MAPT* 10+16 mutation); patient 3 carried a non-specific *C9orf72* mutation detected via TP-PCR with over 30 hexanucleotide repeats; patient 6 carried a c.388_391delCAGT, p.(Gln130Ser)fs *GRN* mutation.

Brain tissues from five age- and sex-matched controls (Table 1) were included for group comparisons with patients at post-mortem. Controls were defined by the absence of significant neurodegenerative pathology at autopsy and Braak tangle stages ≤ III.

### Neuroimaging protocol and processing

Participants with FTD underwent *in vivo* TSPO PET scans with [¹¹C]PK11195 tracer, and structural MRI. MRI used Siemens Magnetom Tim Trio and Verio scanners (Siemens Healthineers, Erlangen, Germany) with an MPRAGE T1-weighted sequence, while PET used GE Advance and GE Discovery 690 PET/CT (GE Healthcare, Waukesha, USA) scanners. MRI and PET data were acquired and processed using previously described methods (Bevan-Jones et al. 2017, Malpetti et al 2023 Brain). Briefly, patients underwent a 3T MRI scan, followed by a dynamic [¹¹C]PK11195 PET scan for 75 minutes. Individual PET images were rigidly co-registered to the T-1 MRI scans. The tissue time-activity curve was determined by supervised cluster analysis. Non-displaceable binding potential (BPND) was calculated in cortical (Brodmann areas) and subcortical (modified n30r83 Hammersmith atlas, including brainstem parcellation and the cerebellar dentate nucleus) regions using a simplified tissue reference model with correction for vascular binding (Yaqub et al., 2012; Malpetti et al., 2020)

### Brain tissue sampling and preparation

The death-to-formalin post-mortem interval was 64.1 ± 12.4 h in individuals with FTD (median 64, range 39-83) and 57.4 ± 22.3 h in controls (median 51, range 32-92); the groups did not differ (Wilcoxon W = 33.5, *P* = 0.327). Tissue was formalin-fixed paraffin-embedded (FFPE) and stored at the Cambridge Brain Bank. FFPE blocks were sampled in 15 μm tissue sections from: Brodmann areas BA1/2/3 (jointly), BA4, BA6, BA7, BA17/18, BA21/22, BA46, amygdala, cingulate, cerebellar cortex, globus pallidus, putamen, hippocampal cortex, thalamus, medulla, midbrain, and pons.

### Immunohistochemistry protocol across all brain regions

Across all selected brain regions of patients and controls, DAB-based immunohistochemistry was performed to quantify CD68+ microglial load, phosphorylated tau (AT8), and TDP-43 aggregate burden (Table S5) with the same protocol as Wijesinghe et al., 2025. Whole-slide images were acquired on an Aperio AT2 whole-slide scanner (Leica) at ×40 magnification. Full-resolution images were processed in Halo (Indica Labs), where a tissue classifier (DenseNet V2, Indica Labs) was trained with manual annotations to differentiate between grey and white matter while simultaneously excluding non-tissue area, staining artefacts, and meningeal tissue. Grey- and white-matter areas were then colourimetrically quantified for CD68+ stain area.

### Immunofluorescence protocol in frontal, temporal and occipital regions

Fluorescent quadruple staining was performed for marker-co-localisation analyses in three frontotemporal cortical regions across all participants: inferior frontal gyrus (BA46) and temporal cortex (BA21/22), as primary affected in FTLD, and motor/premotor cortex (BA6) as a secondary affected region.

In brief, for co-staining fluorescent analysis, the FFPE slices were baked at 60 °C for 120 minutes. Following this, slides were rehydrated and deparaffinized with a series of baths; first washed in Xylene overnight, then moved to a separate tank with Xylene for 10 minutes. Slices were then washed sequentially in 100% Ethanol, 96% Ethanol, 70% Ethanol, 50% Ethanol, and finally RO H2O for 10 minutes each. Phosphate-buffered saline (PBS) solution is prepared from Tablets (Thermo Fisher, 18912014) in reverse osmosis (RO) H2O. PBS-T is prepared by adding 0.05% Tween® 20 (Sigma-Aldrich) to PBS prepared as described above. The slides then proceeded to antigen retrieval in TRIS-EDTA (pH 9.2) at 90 °C for 20 minutes. After cooling to room temperature, slides were washed in PBS 3 times for 5 minutes per wash. Slides were laid flat outside of the solution and contoured with an Advanced hydrophobic PAP pen (Z672548, Sigma Aldrich), then blocked with 2.5% Bovine Serum Albumin (BSA) in PBS-T for 60 minutes, after which primary antibody incubation was performed overnight with 2.5% BSA in PBS-T. On the following day, slides were washed in PBS 3 times for 5 minutes per wash. After which, Biotium TrueBlack® Lipofuscin Autofluorescence Quencher (A11055, Invitrogen) in 70% ethanol was applied to the slides for auto-fluorescent quenching for 30 seconds for the IBA1 stains and 45 seconds for the GFAP and CD31 stains. Subsequently, the slides were washed in PBS for 5 minutes 2 times. Slides were incubated with the appropriate secondary antibodies (See Table S6) in 2.5% BSA in PBS for 120 minutes. Following this incubation, slides were washed in PBS 3 times for 5 minutes each. Nucleic acids were stained with DAPI – 4’,6-diamidino-2-phenylindole, dihydrochloride– (ThermoFisher, 62247) immediately after for 10 minutes. Finally, slides were washed in PBS 3 times for 5 minutes per wash and then coverslipped (Corning®, 2980-245) with Fluoromount-G™ Mounting Medium (Invitrogen, 00-4958-02).

The protocol used two vicinal slide sets per subject and region, prepared from sequential sections of the same FFPE block: **Set 1** (DAPI / IBA1 / CD68 / TSPO) and **Set 2** (DAPI / GFAP / CD31 / TSPO). The TSPO antibody, dilution, and image-acquisition channel were the same across both slide sets. TrueBlack® Lipofuscin Autofluorescence Quencher (Biotium) was used to minimise tissue autofluorescence. Primary and secondary antibodies, with dilutions and antigen retrieval buffers, are listed in Tables S5 and S6 respectively.

Fluorescent image acquisition for the co-localisation analysis was performed on a Zeiss Axioscan Z1 Slidescanner at 40×. For the single-cell co-localisation experiment, a Leica Stellaris 8 confocal microscope (40× water-immersion objective (HC PL APO 40x/1.10 W); FOV 228.16 × 228.16 μm) was used in Lightning™ deconvolution mode, with system-optimised Z-stacks acquired per image (depth ∼10 μm; step ∼0.36 μm). For quantification, six sub-images were extracted per slice and processed in ImageJ (Fiji) using custom BIOP JaCoP macros: background subtraction (rolling ball radius = 13 μm, sliding paraboloid, no smoothing), Gaussian blur (σ = 2 μm, scaled units), and contrast enhancement (0.35% saturation) per channel. Co-localisation analyses used the BIOP JaCoP plug-in with Triangle thresholds applied to cell-marker channels (IBA1, CD68, GFAP) and Otsu thresholds applied to the TSPO channel. For the CD31-TSPO dataset, the four-channel images were first split to allow specific preprocessing of channel 4 (CD31): channels 1-3 were processed as above, while channel 4 underwent additional filtering (rolling-ball background subtraction radius = 120 μm, median filter radius = 2 μm, Gaussian blur σ = 2 μm, and local-maxima enhancement radius = 13 μm). The channels were then re-merged prior to co-localisation analysis, computed between CD31 and TSPO using Moments and Otsu thresholds respectively.

### Three-dimensional single-cell quantification

Three-dimensional confocal image stacks were processed in Imaris 11 (Andor Oxford Instruments) to quantify intracellular TSPO and CD68 content at the level of individual IBA1+ microglia and CD68+ cells. Prior to object detection, each channel was preprocessed independently: a Gaussian filter (σ = 0.223 μm) was applied to the IBA1 channel; 3D median filters (kernel 3×3×1 voxels) were applied to the TSPO and CD68 channels, with an additional background subtraction step applied to the CD68 channel (σ = 57.088 μm).

For the IBA1-based pipeline, microglial cell bodies were segmented as 3D surfaces using a classifier trained on all four imaging channels, with IBA1 fluorescence as the primary reference. Surface grain size was set to 0.446 μm and the diameter of the largest sphere to 1.67 μm. Cell bodies were retained based on minimum voxel volume (≥ 6,333 voxels) and nucleus sphericity (≥ 0.46). TSPO+ and CD68+ puncta were detected as vesicle objects within each segmented cell body using local-contrast thresholding (estimated diameters: TSPO 1.0 μm isotropic; CD68 2.23 μm). The intracellular TSPO fraction and intracellular CD68 fraction were defined as the volume of vesicle-type objects occupying the cell-marker-positive cell volume, expressed as a proportion of total cell-body volume.

For the CD68-based pipeline, cell bodies were segmented from the CD68 channel alone using a classifier trained on this channel exclusively, with equivalent surface parameters. TSPO+ vesicle objects were detected within the resulting CD68+ cell volumes using an estimated diameter of 1.0 μm and local-contrast thresholding.

### Statistical analyses

Analyses were performed in R 4.5.3. Group descriptives used Wilcoxon rank-sum and Fisher’s exact tests. Inferential models were frequentist linear mixed-effects models (LMEMs) with Kenward-Roger degrees of freedom; marginal means, slope contrasts, and pairwise comparisons used emmeans / emtrends. Single-cell Imaris tests used Wilcoxon rank-sum (exact) with Hodges-Lehmann shifts and 95% confidence intervals. Uncorrected P-values are reported throughout; Benjamini-Hochberg FDR correction was applied where indicated.

Random-effects structure: For LMEMs with a continuous within-subject predictor (TSPO PET BPND), the random-effects structure was selected per fit following Matuschek et al. (2017): three candidates were fit by REML and compared; random intercept only (1|id), random slope only (0+BPND|id), and correlated random intercept plus slope (1+BPND|id). Fits flagged as singular by lme4’s isSingular were discarded; the lowest-AIC non-singular structure was retained. For binary group-only contrasts with no continuous within-subject predictor, the structure was fixed a priori at a subject random intercept.

### Detailed statistics

Analyses were performed in R 4.5.3 (aarch64-apple-darwin20). Packages used: tidyverse 2.0.0 (ggplot2 4.0.2, dplyr 1.2.1, tidyr 1.3.2, readr 2.2.0, purrr 1.2.1, tibble 3.3.1, stringr 1.6.0, forcats 1.0.1), readxl 1.4.5, janitor 2.2.1, lme4 2.0.1, lmerTest 3.2.1, emmeans 2.0.2, performance 0.16.0, ragg 1.5.2, patchwork 1.3.2, scales 1.4.0, Matrix 1.7.4, numDeriv 2016.8.1.1, estimability 1.5.1, mvtnorm 1.3.6, systemfonts 1.3.2, and textshaping 1.0.5.

Regional CD68 burden. CD68 ∼ Group × Region + (1|id), with the Group main effect giving the global FTD-Control contrast and Group×Region testing regional heterogeneity; per-region contrasts via emmeans. A parallel model added Segmentation as a further interaction to localise effects to grey matter, white matter, or full tissue. BA4 was excluded from the FTD-versus-control comparison owing to absent control tissue coverage for this region.

TSPO PET ∼ CD68 burden and tissue TSPO area. CD68 ∼ BPND + PET-to-post-mortem interval + (random effects), with a pathology-subgroup interaction (FTLD-tau vs FTLD-TDP) tested separately. Tissue TSPO area was first modelled per slide set (TSPO ∼ Group + Region + (1|id)), then jointly across slide sets to compare slopes (TSPO ∼ BPND + Slide Set + PET-to-post-mortem interval + (random effects)). Exploratory protein-aggregate associations (tau × CD68 in FTLD-tau; pTDP-43 × CD68 in FTLD-TDP) mirrored the BPND-CD68 structure.

Cell-type TSPO co-localisation. Per-marker LMEMs across BA6, BA46, BA21/22: Coloc ∼ Group + Region + (1|id) for the group contrast, and Coloc ∼ BPND + PET-to-post-mortem interval + (random effects) for the regional gradient, one per marker.

Single-cell Imaris metrics. Per-subject means of per-cell values, mean-of-ratios, GM and WM pooled, in Control (n = 4) and FTD (n = 10). Within-group: for each metric, per-subject CD68− vs CD68+ means were compared by exact paired Wilcoxon signed-rank test, separately in Control and FTD (Fig 5). Between-group (supplementary): per-subject metrics were compared FTD vs Control by exact Wilcoxon rank-sum test with Hodges–Lehmann shifts and 95% CIs (Fig S5). p-values were BH-FDR corrected within each family.

## Data Availability

Anonymized data used for this analysis is available upon reasonable request. Further participant information, images or samples can be requested but are likely to require a data/material transfer agreement to adhere to consent restrictions including protection of confidentiality.

## Acknowledgements

We thank our participant volunteers and their families for their participation in this study and donations to the Cambridge Brain Bank. We thank the National Institute for Health Research (NIHR) Cambridge Biomedical Research Centre, and the research nurses for their contribution. We thank the radiographers and technicians at the Wolfson Brain Imaging Centre and Addenbrooke’s Hospital PET/CT Unit for their role in data acquisition. For the purpose of open access, the authors have applied a Creative Commons Attribution (CC BY) license to any Author Accepted Manuscript version arising from this submission. This work is licensed under a Creative Commons Attribution 4.0 International License.

The authors would like to thank Kristy Halliday and Daniel Rodgers from the Cambridge Brain Bank for their assistance with tissue sectioning and scanning.

## Author’s Contributions

DSV, JBR, MGS and MM contributed to study design. DSV, KOL, MB, GN, DM, JG, NLS, HC, TF, YH, FA, EA, KSJA, JTO, MM contributed to execution, data collection and/or analysis. DSV, KSJA, AQ, JBR, MM contributed to sample selection, preparation and/or staging. DSV, KOL, and MM drafted the first version of the manuscript. DSV, KOL, and MB oversaw the sample and data analysis. MGS and MM supervised the execution and data analysis. DSV, KOL, MB, GN, DCM, AA, JG, NLS, HC, TF, YH, SW, FA, EA, KSJA, AQ, JTO, JBR, MGS, and MM reviewed the manuscript and approved the final version of the manuscript.

## Supplementary Material

### A1. Imaris sub-cohort and pathology-class assignment

The full demographic, clinical and neuropathological cohort information is in **Table 1**. This appendix summarises the pathology-subgroup assignment used in the Tau / TDP analyses (§A2) and the Imaris sub-cohort (§6, Fig. 5).

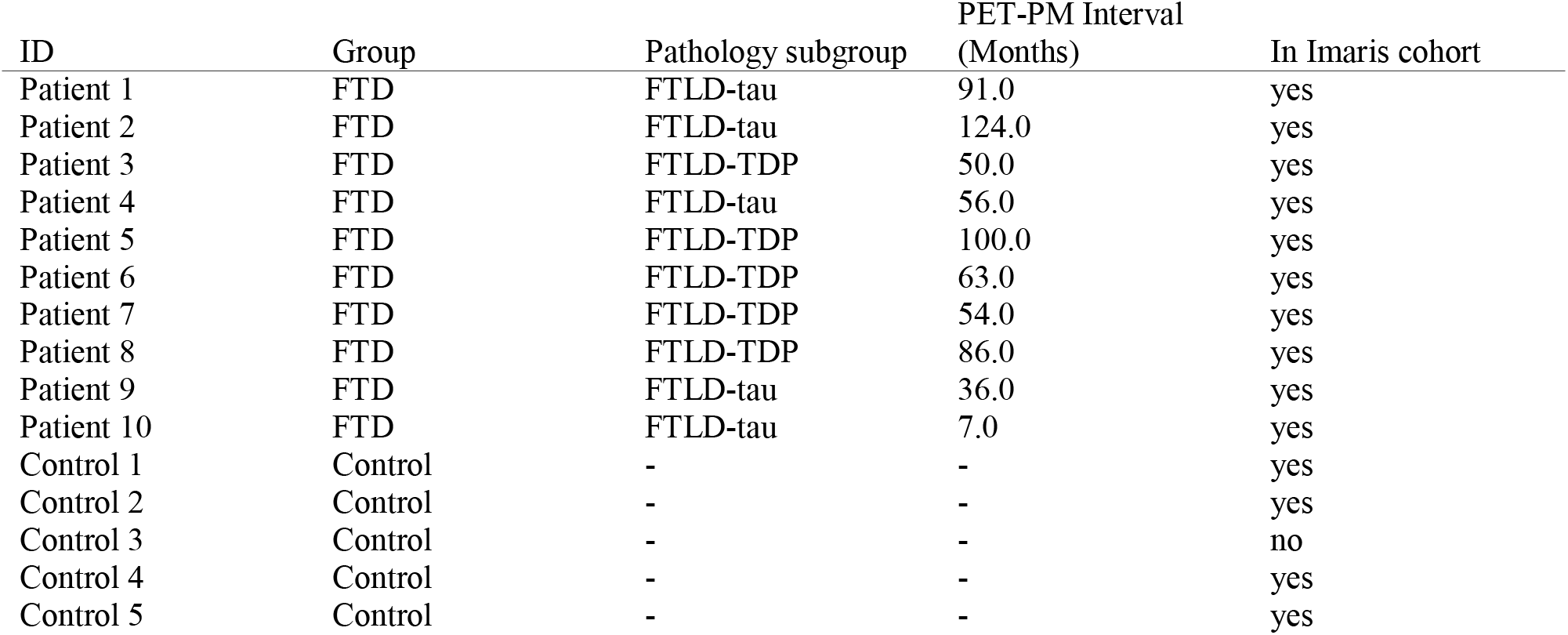

| ID | Group | Pathology subgroup | PET-PM Interval<br>(Months) | In Imaris cohort |
| --- | --- | --- | --- | --- |
| Patient 1 | FTD | FTLD-tau | 91.0 | yes |
| Patient 2 | FTD | FTLD-tau | 124.0 | yes |
| Patient 3 | FTD | FTLD-TDP | 50.0 | yes |
| Patient 4 | FTD | FTLD-tau | 56.0 | yes |
| Patient 5 | FTD | FTLD-TDP | 100.0 | yes |
| Patient 6 | FTD | FTLD-TDP | 63.0 | yes |
| Patient 7 | FTD | FTLD-TDP | 54.0 | yes |
| Patient 8 | FTD | FTLD-TDP | 86.0 | yes |
| Patient 9 | FTD | FTLD-tau | 36.0 | yes |
| Patient 10 | FTD | FTLD-tau | 7.0 | yes |
| Control 1 | Control | - | - | yes |
| Control 2 | Control | - | - | yes |
| Control 3 | Control | - | - | no |
| Control 4 | Control | - | - | yes |
| Control 5 | Control | - | - | yes |

### A2. Aggregate (Tau / pTDP-43) ∼ CD68

In an exploratory analysis, within each FTLD class, protein-aggregate burden tracked microglial load weakly. Each subtype was fitted separately by LMEM (response = aggregate %DAB, fixed predictor = CD68 %; *n* = 5 per group), with random effects chosen by the Matuschek ladder. Only Tau demonstrated a weak correlation with CD68 before FDR correction.

- (A) FTLD-tau — tau %DAB rose modestly with CD68 (LMEM [RI]: β = +0.0552, SE = 0.0265, *t* = 2.09, *P* = 0.039, *P*_FDR = 0.079; R²m = 0.032).
- (B) FTLD-TDP — pTDP-43 %DAB was unrelated to CD68 (LMEM [RI]: β = −0.1074, SE = 0.0905, *t* = −1.19, *P* = 0.241, *P*_FDR = 0.241; R²m = 0.027).

**Figure S1.**
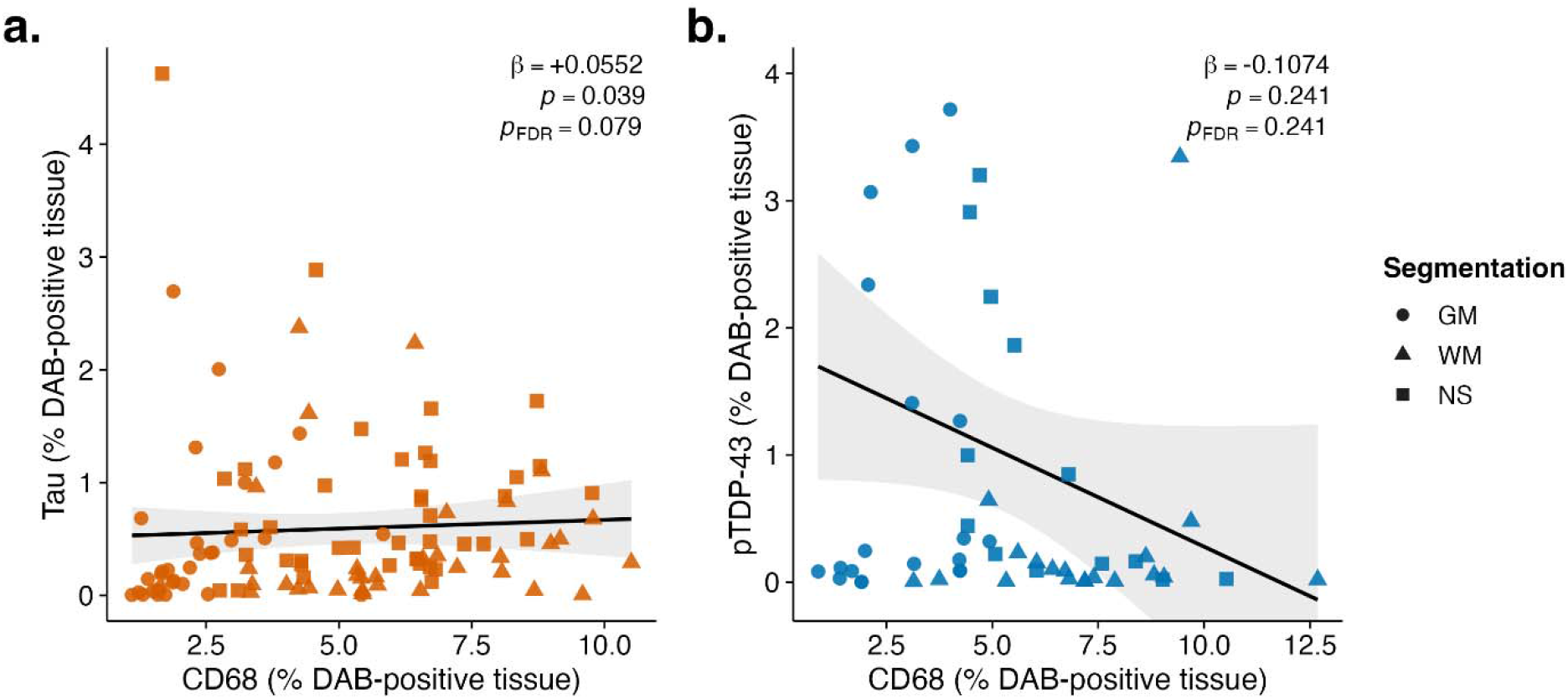
Protein-aggregate burden against CD68 burden across all regions in FTLD-tau patients (A) and FTLD-TDP patients (B), separately. **(A)** Tau %DAB ∼ CD68 in FTLD-tau (n = 5); β = +0.0552, SE = 0.0265, t = 2.09, *P* = 0.039, *P*_FDR = 0.079. **(B)** pTDP-43 %DAB ∼ CD68 in FTLD-TDP (n = 5); β = −0.1074, SE = 0.0905, t = −1.19, *P* = 0.241, *P*_FDR = 0.241. Per-subset LMEM with Matuschek-selected RE (both subsets selected RI).

### A3. Tissue TSPO area fraction × BPND × Segmentation - Figure S2

Splitting tissue into grey and white matter did not improve on the model of §3, so the TSPO area-fraction–BPND relationship holds across both compartments rather than being driven by one. Segmentation main effect (LRT 1-vs-2); not preferred over the BPND-only baseline (χ² = 0.224, df = 1, *P* = 0.636). BPND × Segmentation interaction (LRT 2-vs-3); not preferred over the additive model (χ² = 0.337, df = 1, *P* = 0.562). GM slope (REML refit of rung 3); β = +0.0560, SE = 0.0404, *t* = 1.39, *P* = 0.168, *P*_FDR = 0.233 (Benjamini–Hochberg within the 2-slope family). WM slope — β = +0.0303, SE = 0.0252, *t* = 1.20, *P* = 0.233, *P*_FDR = 0.233. GM − WM contrast — slopes look distinct but do not differ statistically (+0.0257, SE = 0.0482, *t* = 0.53, *P* = 0.595).

See also Fig. S3 for the per-marker (CD68 / IBA1 / GFAP / CD31) segmentation cascade on the marker-TSPO co-localised area.

**Figure S2.**
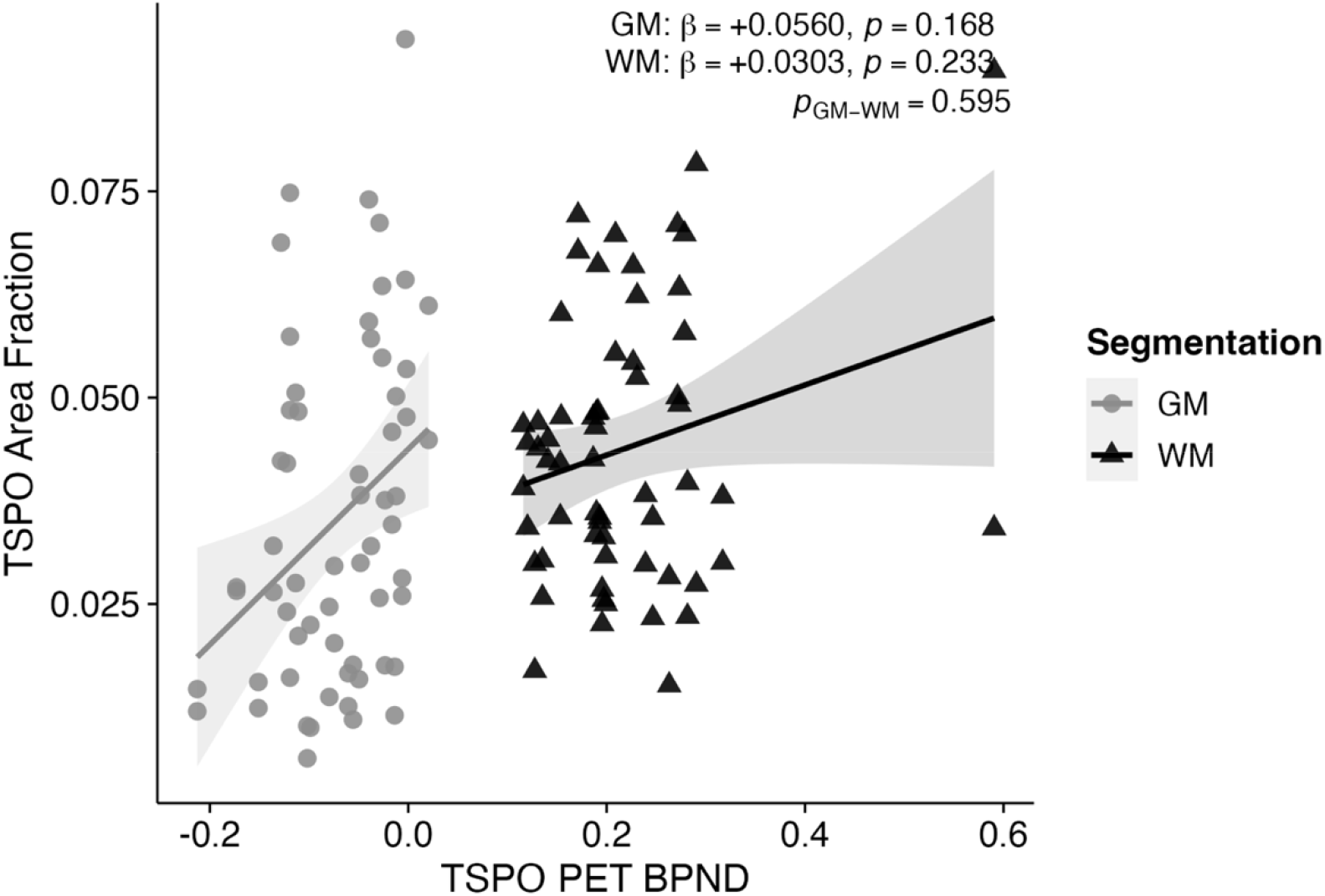
Tissue TSPO area fraction × BPND, decomposed by tissue segmentation. Tissue TSPO area fraction modelled by LMEM (*Post mortem TSPO ∼ TSPO PET * Segmentation + SlideSet + PET_PM_Interval + (1|id)*) within the 10 patients with FTD. Per-Segmentation BPND slopes: GM β = +0.0560 (SE = 0.0404, t = 1.39, P = 0.168); WM β = +0.0303 (SE = 0.0252, t = 1.20, P = 0.233). The grey-versus white-matter slope contrast is +0.0257 (SE = 0.0482, t = 0.53, P = 0.595), i.e. the BPND–area-fraction association does not differ between compartments. Supporting nested-model tests: Segmentation main effect χ² = 0.224, df = 1, P = 0.636; BPND × Segmentation χ² = 0.337, df = 1, P = 0.562.

### A4. Coloc × Segmentation - GM-driven decomposition - Figure S3

A grey-matter / white-matter decomposition (Supplementary Figure S3) shows the CD68 positive coupling concentrated in grey matter (GM β = +0.013, P = 0.052; WM β = −0.009, P = 0.027; TSPO PET signal × Segmentation interaction LRT P = 0.007; GM–WM slope contrast P = 0.006, P_FDR = 0.012), with the same compartment-specific structure for IBA1 (GM β = +0.048, P = 0.006; WM β = −0.016, P = 0.122; LRT P = 0.001; contrast P = 0.002, P_FDR = 0.008). GFAP and CD31 show no significant compartmental difference (LRT P = 0.310 and 0.88; contrast P = 0.281 and 0.951; P_FDR = 0.375 and 0.951).

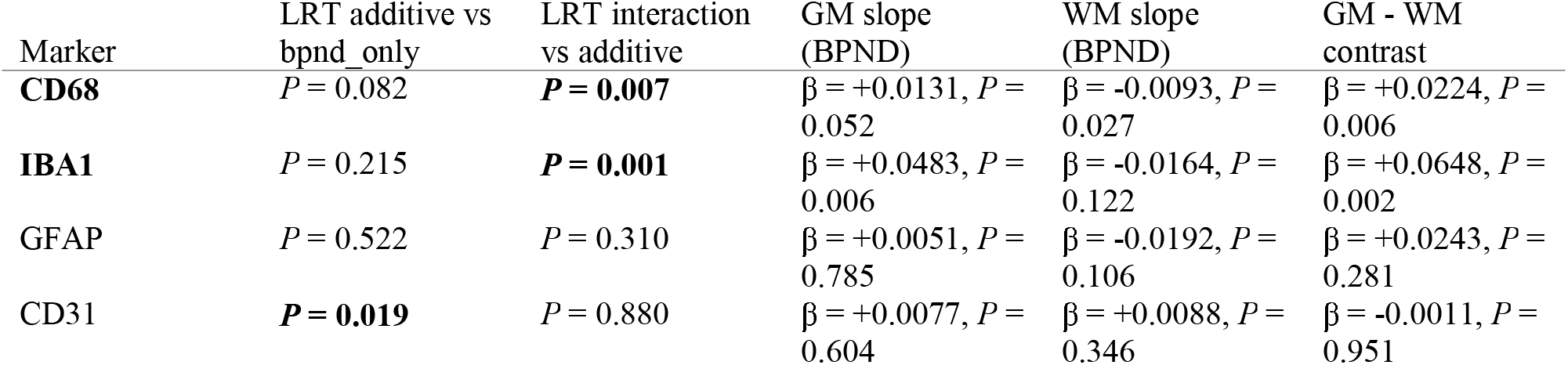

**Figure S3.**
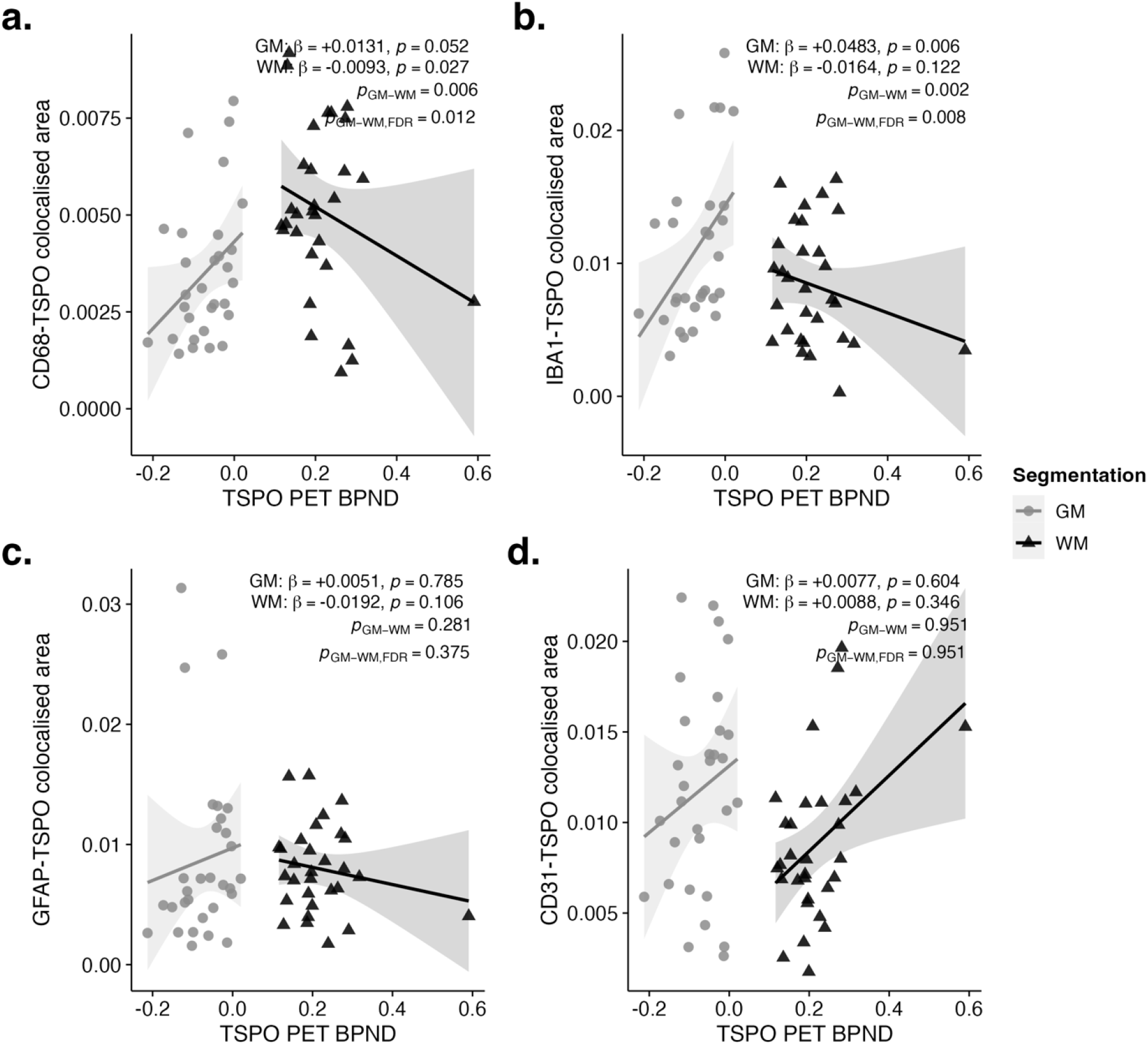
Marker–TSPO co-localised area × BPND, decomposed by tissue segmentation. Marker–TSPO co-localised area modelled by LMEM with regional TSPO PET binding (BPND) as the fixed-effect predictor, within all 10 patients with FTD, decomposed by grey and white matter (*Coloc_by_Total ∼ BPND * Segmentation + PET_PM_Interval + (1|id)*). (A) CD68. (B) IBA1. (C) GFAP. (D) CD31. Per-segmentation points and linear-fit slope ribbons are coloured GM = grey (filled circles), WM = black (filled triangles). Per-compartment BPND slopes (β) with their p-values are estimated marginal trends (emtrends). pGM–WM is the model-based test of the grey-versus white-matter BPND-slope difference (the BPND × Segmentation contrast); pGM–WM,FDR is its Benjamini–Hochberg-adjusted value across the four markers (family m = 4). The CD68 and IBA1 slopes differ significantly between compartments (pGM–WM,FDR = 0.012 and 0.008), with positive GM and negative WM coupling; GFAP and CD31 show no compartmental difference (pGM–WM,FDR = 0.375 and 0.951).

### A5. Cell marker tissue-fraction companion to §5

In the same slides as §5 CD68 and IBA1 co-localised marker area diverge in direction across regions while GFAP and CD31 did not, reproducing the §5 pattern. Random intercept (Matuschek-selected; Marker Area ∼ TSPO PET + PET_PM_Interval + (1|id)) for all four markers, with BH correction.

- (A) CD68 — β_BPND = +0.0465, SE = 0.0089, *t* = 5.25, *P* < 0.001, *P*_FDR < 0.001.
- (B) IBA1 — β_BPND = −0.0720, SE = 0.0156, *t* = −4.62, *P* < 0.001, *P*_FDR < 0.001.
- (C) GFAP — β_BPND = −0.1394, SE = 0.0463, *t* = −3.01, *P* = 0.004, *P*_FDR = 0.005.
- (D) CD31 — β_BPND = −0.0209, SE = 0.0076, *t* = −2.74, *P* = 0.008, *P*_FDR = 0.008.

**Figure S4.**
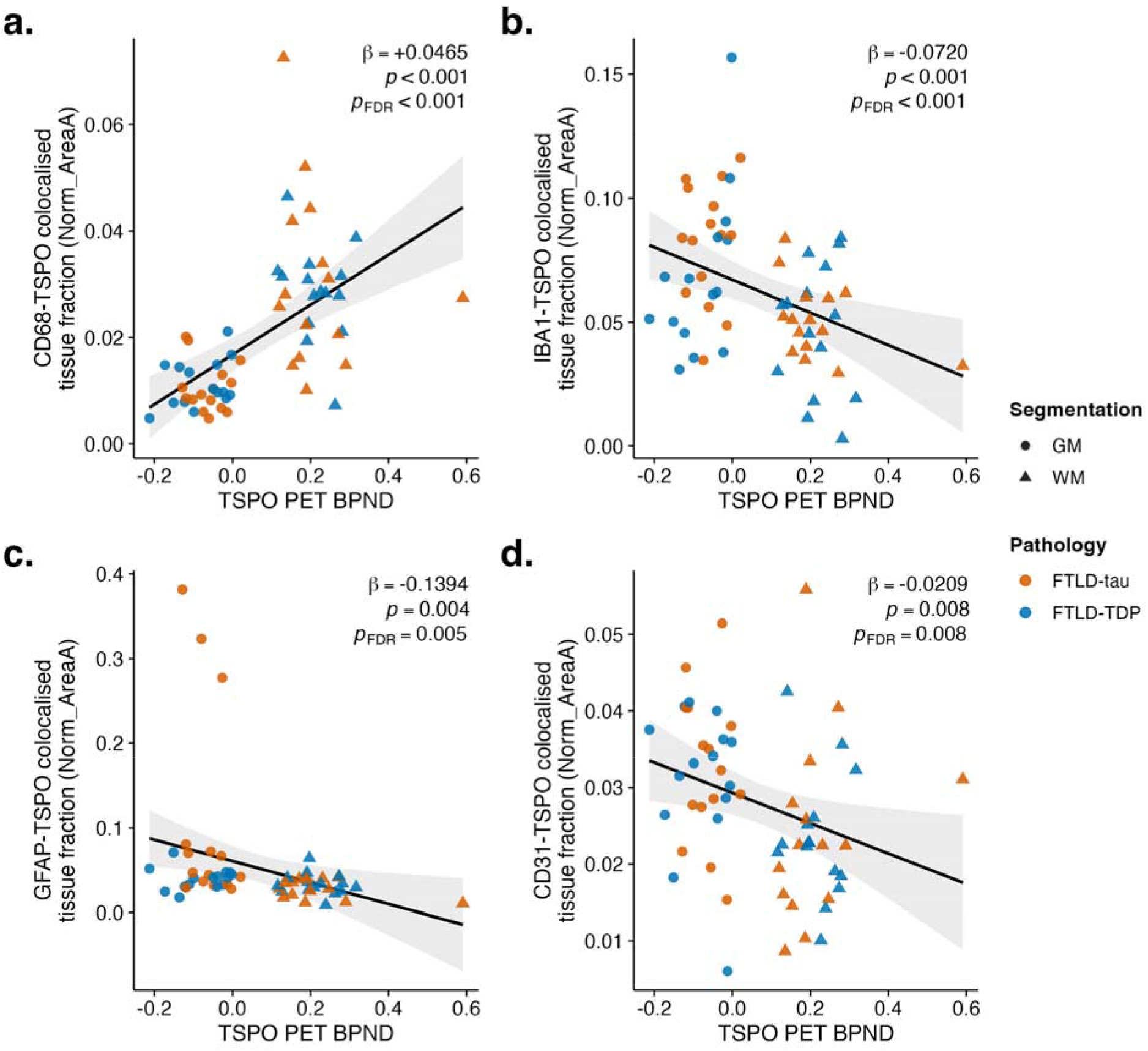
Cell marker tissue fraction × TSPO PET, four-marker companion. Marker area expressed as a fraction of total tissue modelled by LMEM, fitted per marker. LMEMs Matuschek-selected; Marker Area ∼ TSPO PET + PET_PM_Interval + (1|id) **(A)** CD68: β_BPND = +0.0465, SE = 0.0089, t = 5.25, *P* < 0.001, *P*_FDR < 0.001. **(B)** IBA1: β_BPND = −0.0720, SE = 0.0156, t = −4.62, *P* < 0.001, *P*_FDR < 0.001. **(C)** GFAP: β_BPND = −0.1394, SE = 0.0463, t = −3.01, *P* = 0.004, *P*_FDR = 0.005. **(D)** CD31: β_BPND = −0.0209, SE = 0.0076, t = −2.74, *P* = 0.008, *P*_FDR = 0.008. RE = RI selected by Matuschek for all four markers.

**Figure S5.**
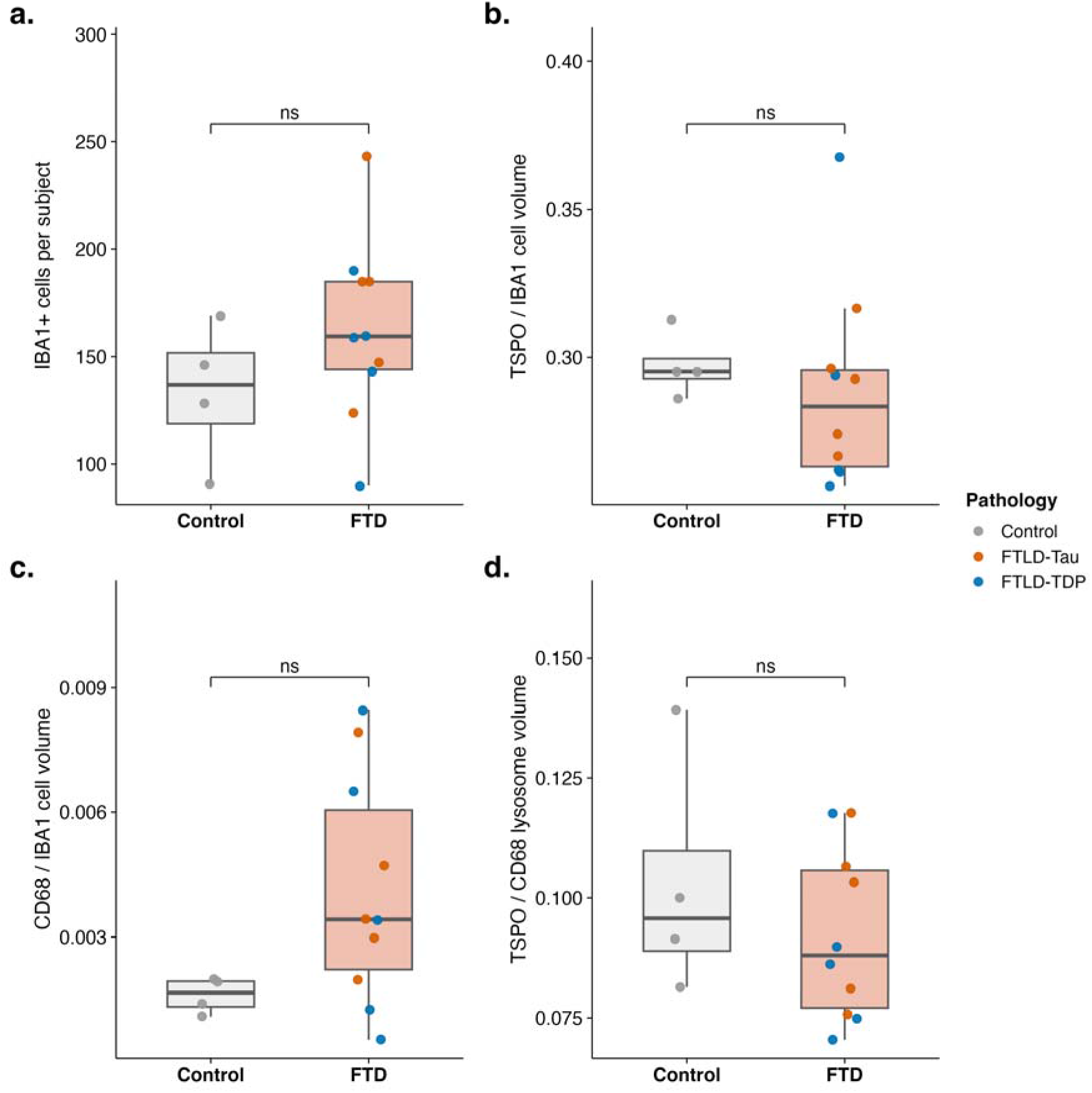
Subject-level single-cell Imaris metrics do not differ between groups. Colour palette: Control = grey, FTLD-tau = orange, FTLD-TDP = blue; box fill salmon = FTD, light grey = Control; points coloured by pathology subgroup. Per-subject values compared between FTD and Control by Wilcoxon rank-sum test (grey and white matter pooled), with Hodges–Lehmann shift and 95% CI; BH-FDR across the four-test family (m = 4). (A) IBA1+ cell density per subject; W = 28, P = 0.288, P_FDR = 0.454, HL = +25.6 [−22, +94]. (B) Intracellular TSPO volume fraction in IBA1+ microglia; W = 13, P = 0.374, P_FDR = 0.454, HL = −0.019 [−0.039, +0.021]. (C) Intracellular CD68 volume fraction in IBA1+ microglia; W = 32, P = 0.106, P_FDR = 0.424, HL = +0.0019 [−0.0006, +0.0065]. (D) Intracellular TSPO volume fraction within CD68+ cells; W = 14, P = 0.454, P_FDR = 0.454, HL = −0.010 [−0.049, +0.022].

### A6. Antibody tables (primary and secondary)

**Table S5.**
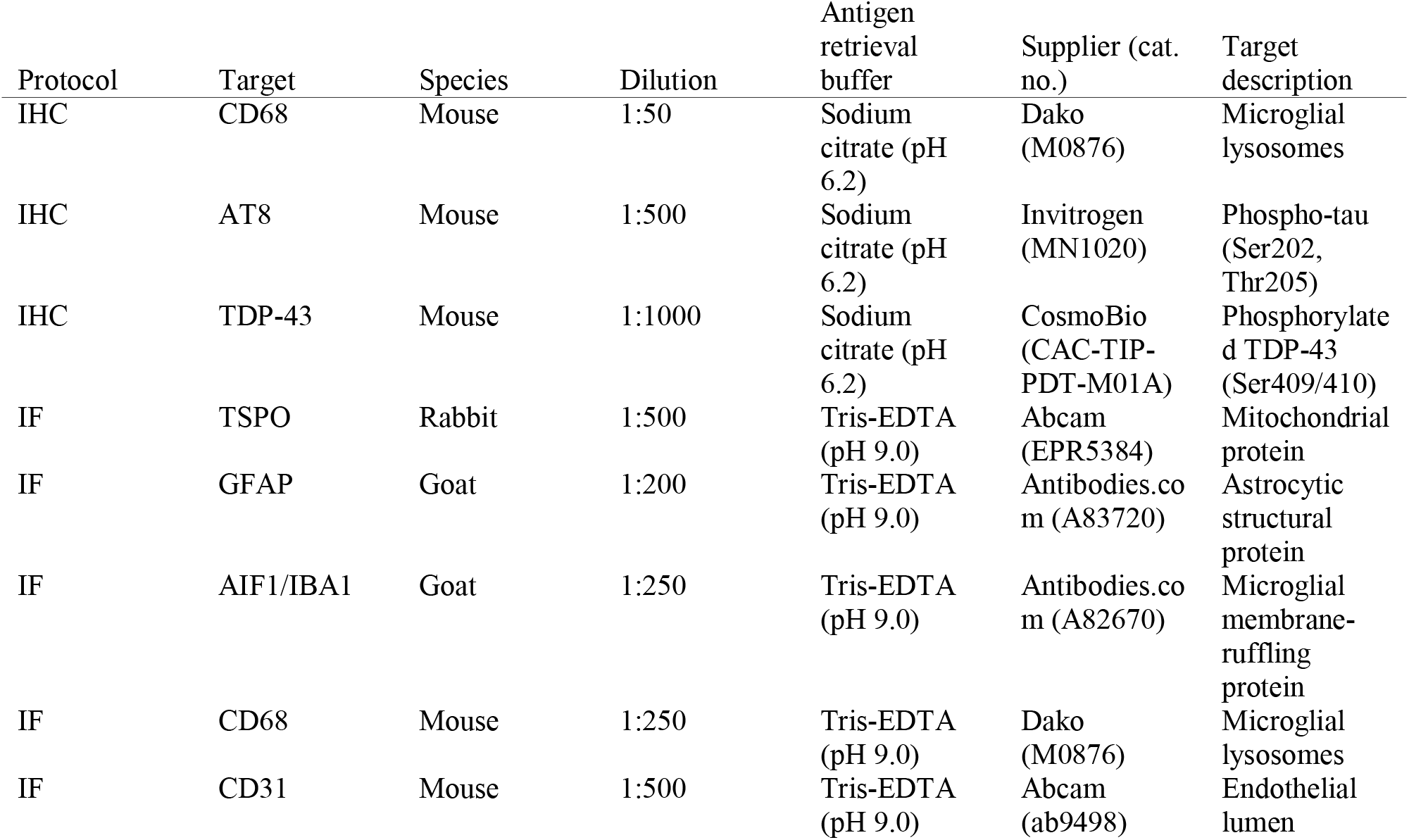
Primary antibodies used for immunohistochemistry (IHC) and immunofluorescence (IF).

**Table S6.** Secondary antibodies.

| Protocol | Target species | Host | Dilution | Fluorophore | Supplier (cat. no.) |
| --- | --- | --- | --- | --- | --- |
| IHC | Mouse | Donkey | 1:500 | n/a (HRP) | ThermoFisher (A24506) |
| IF | Rabbit | Donkey | 1:500 | AF568 | ThermoFisher (A10042) |
| IF | Goat | Donkey | 1:500 | AF488 | ThermoFisher (A-11055) |
| IF | Mouse | Donkey | 1:500 | AF647 | ThermoFisher (A-31571) |

## References

Alster, Piotr, and Natalia Madetko-Alster. 2025. “The Genetic Background of the Immunological and Inflammatory Aspects of Progressive Supranuclear Palsy.” International Journal of Molecular Sciences 26(9): 3927. doi:10.3390/ijms26093927.

Appleton, Johanna, Quentin Finn, Paolo Zanotti-Fregonara, Meixiang Yu, Alireza Faridar, Mohammad O Nakawah, Carlos Zarate, et al. 2025. “Brain Inflammation Co-Localizes Highly with Tau in Mild Cognitive Impairment Due to Early-Onset Alzheimer’s Disease.” Brain 148(1): 119–32. doi:10.1093/brain/awae234.

Bevan-Jones, W Richard, Thomas E Cope, P Simon Jones, Sanne S Kaalund, Luca Passamonti, Kieren Allinson, Oliver Green, et al. 2020. “Neuroinflammation and Protein Aggregation Co-Localize across the Frontotemporal Dementia Spectrum.” Brain 143(3): 1010–26. doi:10.1093/brain/awaa033.

Bevan-Jones, W Richard, Ajenthan Surendranathan, Luca Passamonti, Patricia Vázquez Rodríguez, Robert Arnold, Elijah Mak, Li Su, et al. 2017. “Neuroimaging of Inflammation in Memory and Related Other Disorders (NIMROD) Study Protocol: A Deep Phenotyping Cohort Study of the Role of Brain Inflammation in Dementia, Depression and Other Neurological Illnesses.” BMJ Open 7(1): e013187. doi:10.1136/bmjopen-2016-013187.

Bright, Fiona, Eryn L. Werry, Carol Dobson-Stone, Olivier Piguet, Lars M. Ittner, Glenda M. Halliday, John R. Hodges, et al. 2019. “Neuroinflammation in Frontotemporal Dementia.” Nature Reviews Neurology 15(9): 540–55. doi:10.1038/s41582-019-0231-z.

Cosenza Nashat, M., M. L. Zhao, H. S. Suh, J. Morgan, R. Natividad, S. Morgello, and S. C. Lee. 2009. “Expression of the Translocator Protein of 18 kDa by Microglia, Macrophages and Astrocytes Based on Immunohistochemical Localization in Abnormal Human Brain.” Neuropathology and Applied Neurobiology 35(3): 306–28. doi:10.1111/j.1365-2990.2008.01006.x.

De Picker, Livia J., Manuel Morrens, Igor Branchi, Bartholomeus C.M. Haarman, Tatsuhiro Terada, Min Su Kang, Delphine Boche, et al. 2023. “TSPO PET Brain Inflammation Imaging: A Transdiagnostic Systematic Review and Meta-Analysis of 156 Case-Control Studies.” Brain, Behavior, and Immunity 113: 415–31. doi:10.1016/j.bbi.2023.07.023.

Doorn, Karlijn J, Tim Moors, Benjamin Drukarch, Wilma D.J. van de Berg, Paul J Lucassen, and Anne-Marie van Dam. 2014. “Microglial Phenotypes and Toll-like Receptor 2 in the Substantia Nigra and Hippocampus of Incidental Lewy Body Disease Cases and Parkinson’s Disease Patients.” Acta Neuropathologica Communications 2(1): 90. doi:10.1186/s40478-014-0090-1.

Fairley, Lauren H., Kei Onn Lai, Amandine Grimm, Anne Eckert, and Anna M. Barron. 2024. “The Mitochondrial Translocator Protein (TSPO) in Alzheimer’s Disease: Therapeutic and Immunomodulatory Functions.” Biochimie 224: 120–31. doi:10.1016/j.biochi.2024.07.003.

Fan, Zhen, Aren A. Okello, David J. Brooks, and Paul Edison. 2015. “Longitudinal Influence of Microglial Activation and Amyloid on Neuronal Function in Alzheimer’s Disease.” Brain 138(12): 3685–98. doi:10.1093/brain/awv288.

Garland, Emma F., Henrike Antony, Laura Kulagowska, Thomas Scott, Charlotte Rogien, Michel Bottlaender, James A. R. Nicoll, and Delphine Boche. 2024. “The Microglial Translocator Protein (TSPO) in Alzheimer’s Disease Reflects a Phagocytic Phenotype.” Acta Neuropathologica 148(1): 62. doi:10.1007/s00401-024-02822-x.

Garland, Emma F., Oliver Dennett, Laurie C. Lau, David S. Chatelet, Michel Bottlaender, James A. R. Nicoll, and Delphine Boche. 2023. “The Mitochondrial Protein TSPO in Alzheimer’s Disease: Relation to the Severity of AD Pathology and the Neuroinflammatory Environment.” Journal of Neuroinflammation 20(1): 186. doi:10.1186/s12974-023-02869-9.

Gorno-Tempini, M.L., A.E. Hillis, S. Weintraub, A. Kertesz, M. Mendez, S.F. Cappa, J.M. Ogar, et al. 2011. “Classification of Primary Progressive Aphasia and Its Variants.” Neurology 76(11): 1006–14. doi:10.1212/WNL.0b013e31821103e6.

Grossman, Murray, William W. Seeley, Adam L. Boxer, Argye E. Hillis, David S. Knopman, Peter A. Ljubenov, Bruce Miller, et al. 2023. “Frontotemporal Lobar Degeneration.” Nature Reviews Disease Primers 9(1): 40. doi:10.1038/s41572-023-00447-0.

Guilarte, Tomás R., Alexander N. Rodichkin, Jennifer L. McGlothan, Arlet Maria Acanda De La Rocha, and Diana J. Azzam. 2022. “Imaging Neuroinflammation with TSPO: A New Perspective on the Cellular Sources and Subcellular Localization.” Pharmacology & Therapeutics 234: 108048. doi:10.1016/j.pharmthera.2021.108048.

Hendrickx, Debbie A.E., Corbert G. Van Eden, Karianne G. Schuurman, Jörg Hamann, and Inge Huitinga. 2017. “Staining of HLA-DR, Iba1 and CD68 in Human Microglia Reveals Partially Overlapping Expression Depending on Cellular Morphology and Pathology.” Journal of Neuroimmunology 309: 12–22. doi:10.1016/j.jneuroim.2017.04.007.

Hopperton, K E, D Mohammad, M O Trépanier, V Giuliano, and R P Bazinet. 2018. “Markers of Microglia in Post-Mortem Brain Samples from Patients with Alzheimer’s Disease: A Systematic Review.” Molecular Psychiatry 23(2): 177–98. doi:10.1038/mp.2017.246.

Ishizawa, Keisuke, and Dennis W. Dickson. 2001. “Microglial Activation Parallels System Degeneration in Progressive Supranuclear Palsy and Corticobasal Degeneration.” Journal of Neuropathology & Experimental Neurology 60(6): 647–57. doi:10.1093/jnen/60.6.647.

Jensen, Per, Brice Ozenne, Per Meden, Ling Feng, Gerda Thomsen, Lars Knudsen, Henrik Steglich Arnholm, et al. 2025. “Poststroke Translocator Protein Expression Dynamics and Correlations to Chronic Infarction: A^[123I]^ CLINDE SPECT Study.” Journal of Neuroimaging 35(1): e70002. doi:10.1111/jon.70002.

Kim, Min-Jeong, Meghan McGwier, Kimberly J. Jenko, Joseph Snow, Cheryl Morse, Sami S. Zoghbi, Victor W. Pike, Robert B. Innis, and William C. Kreisl. 2019. “Neuroinflammation in Frontotemporal Lobar Degeneration Revealed by^11^ C PBR28 PET.” Annals of Clinical and Translational Neurology 6(7): 1327–31. doi:10.1002/acn3.50802.

Kreisl, William C., Chul Hyoung Lyoo, Jeih-San Liow, Monica Wei, Joseph Snow, Emily Page, Kimberly J. Jenko, et al. 2016. “11C-PBR28 Binding to Translocator Protein Increases with Progression of Alzheimer’s Disease.” Neurobiology of Aging 44: 53–61. doi:10.1016/j.neurobiolaging.2016.04.011.

Malpetti, Maura, Thomas E Cope, Duncan Street, P Simon Jones, Frank H Hezemans, Elijah Mak, Kamen A Tsvetanov, et al. 2023. “Microglial Activation in the Frontal Cortex Predicts Cognitive Decline in Frontotemporal Dementia.” Brain 146(8): 3221–31. doi:10.1093/brain/awad078.

Malpetti, Maura, Luca Passamonti, Peter Simon Jones, Duncan Street, Timothy Rittman, Timothy D Fryer, Young T Hong, et al. 2021. “Neuroinflammation Predicts Disease Progression in Progressive Supranuclear Palsy.” *Journal of Neurology*, Neurosurgery & Psychiatry 92(7): 769–75. doi:10.1136/jnnp-2020-325549.

Malpetti, Maura, Luca Passamonti, Timothy Rittman, P. Simon Jones, Patricia Vázquez Rodríguez, W. Richard Bevan Jones, Young T. Hong, et al. 2020. “Neuroinflammation and Tau Colocalize in Vivo in Progressive Supranuclear Palsy.” Annals of Neurology 88(6): 1194–1204. doi:10.1002/ana.25911.

Malpetti, Maura, Sebastian N. Roemer, Stefanie Harris, Mattes Gross, Johannes Gnörich, Andrew Stephens, Anna Dewenter, et al. 2024. “Neuroinflammation Parallels 18F PI 2620 Positron Emission Tomography Patterns in Primary 4 Repeat Tauopathies.” Movement Disorders 39(9): 1480–92. doi:10.1002/mds.29924.

Masuda, Takahiro, Roman Sankowski, Ori Staszewski, Chotima Böttcher, Lukas Amann, Christian Scheiwe, Stefan Nessler, et al. 2019. “Spatial and Temporal Heterogeneity of Mouse and Human Microglia at Single-Cell Resolution.” Nature 566(7744): 388–92. doi:10.1038/s41586-019-0924-x.

Matuschek, Hannes, Reinhold Kliegl, Shravan Vasishth, Harald Baayen, and Douglas Bates. 2017. “Balancing Type I Error and Power in Linear Mixed Models.” Journal of Memory and Language 94: 305–15. doi:10.1016/j.jml.2017.01.001.

MRC CFAS, Thais Minett, John Classey, Fiona E. Matthews, Marie Fahrenhold, Mariko Taga, Carol Brayne, et al. 2016. “Microglial Immunophenotype in Dementia with Alzheimer’s Pathology.” Journal of Neuroinflammation 13(1): 135. doi:10.1186/s12974-016-0601-z.

Mrdjen, Dunja, Anto Pavlovic, Felix J. Hartmann, Bettina Schreiner, Sebastian G. Utz, Brian P. Leung, Iva Lelios, et al. 2018. “High-Dimensional Single-Cell Mapping of Central Nervous System Immune Cells Reveals Distinct Myeloid Subsets in Health, Aging, and Disease.” Immunity 48(2): 380–395.e6. doi:10.1016/j.immuni.2018.01.011.

Neumann, Manuela, Edward B. Lee, and Ian R. Mackenzie. 2021. “Frontotemporal Lobar Degeneration TDP-43-Immunoreactive Pathological Subtypes: Clinical and Mechanistic Significance.” In *Frontotemporal Dementias*, Advances in Experimental Medicine and Biology, eds. Bernardino Ghetti, Emanuele Buratti, Bradley Boeve, and Rosa Rademakers. Cham: Springer International Publishing, 201–17. doi:10.1007/978-3-030-51140-1_13.

Nutma, Erik, Kelly Ceyzériat, Sandra Amor, Stergios Tsartsalis, Philippe Millet, David R. Owen, Vassilios Papadopoulos, and Benjamin B. Tournier. 2021. “Cellular Sources of TSPO Expression in Healthy and Diseased Brain.” European Journal of Nuclear Medicine and Molecular Imaging 49(1): 146–63. doi:10.1007/s00259-020-05166-2.

Nutma, Erik, Nurun Fancy, Maria Weinert, Stergios Tsartsalis, Manuel C. Marzin, Robert C. J. Muirhead, Irene Falk, et al. 2023. “Translocator Protein Is a Marker of Activated Microglia in Rodent Models but Not Human Neurodegenerative Diseases.” Nature Communications 14(1): 5247. doi:10.1038/s41467-023-40937-z.

Nylund, Marjo, Jussi Lehto, Markus Matilainen, Johan Rajander, Saara Wahlroos, Marcus Sucksdorff, Tanja Kuhlmann, and Laura Airas. 2025. “Longitudinal Accumulation of Glial Activation Measured by TSPO-PET Predicts Later Brain Atrophy in Multiple Sclerosis.” Journal of Neuroinflammation 22(1): 200. doi:10.1186/s12974-025-03519-y.

Palleis, Carla, Julia Sauerbeck, Leonie Beyer, Stefanie Harris, Julia Schmitt, Estrella Morenas Rodriguez, Anika Finze, et al. 2021. “In Vivo Assessment of Neuroinflammation in 4 REPEAT Tauopathies.” Movement Disorders 36(4): 883–94. doi:10.1002/mds.28395.

Paolicelli, Rosa C., Amanda Sierra, Beth Stevens, Marie-Eve Tremblay, Adriano Aguzzi, Bahareh Ajami, Ido Amit, et al. 2022. “Microglia States and Nomenclature: A Field at Its Crossroads.” Neuron 110(21): 3458–83. doi:10.1016/j.neuron.2022.10.020.

Passamonti, Luca, Patricia Vázquez Rodríguez, Young T. Hong, Kieren S.J. Allinson, W. Richard Bevan-Jones, David Williamson, P. Simon Jones, et al. 2018. “[^11^ C]PK11195 Binding in Alzheimer Disease and Progressive Supranuclear Palsy.” Neurology 90(22). doi:10.1212/WNL.0000000000005610.

Quick, Joseph D., Cristian Silva, Jia Hui Wong, Kah Leong Lim, Richard Reynolds, Anna M. Barron, Jialiu Zeng, and Chih Hung Lo. 2023. “Lysosomal Acidification Dysfunction in Microglia: An Emerging Pathogenic Mechanism of Neuroinflammation and Neurodegeneration.” Journal of Neuroinflammation 20(1): 185. doi:10.1186/s12974-023-02866-y.

Rascovsky, Katya, John R. Hodges, David Knopman, Mario F. Mendez, Joel H. Kramer, John Neuhaus, John C. Van Swieten, et al. 2011. “Sensitivity of Revised Diagnostic Criteria for the Behavioural Variant of Frontotemporal Dementia.” Brain 134(9): 2456–77. doi:10.1093/brain/awr179.

Rizzo, Gaia, Mattia Veronese, Matteo Tonietto, Benedetta Bodini, Bruno Stankoff, Catriona Wimberley, Sonia Lavisse, et al. 2019. “Generalization of Endothelial Modelling of TSPO PET Imaging: Considerations on Tracer Affinities.” Journal of Cerebral Blood Flow & Metabolism 39(5): 874–85. doi:10.1177/0271678X17742004.

Shapiro, Noah L, Peter Simon Jones, Elijah Mak, Kamen A Tsvetanov, Julia Goddard, Davi S Vontobel, Robert Durcan, et al. 2025. “Inflammation PET and Plasma Neurofilament Light Predict Survival in People with Progressive Supranuclear Palsy.” Brain Communications 7(6): fcaf467. doi:10.1093/braincomms/fcaf467.

Swanson, Molly E. V., Miran Mrkela, Helen C. Murray, Maize C. Cao, Clinton Turner, Maurice A. Curtis, Richard L. M. Faull, Adam K. Walker, and Emma L. Scotter. 2023. “Microglial CD68 and L-Ferritin Upregulation in Response to Phosphorylated-TDP-43 Pathology in the Amyotrophic Lateral Sclerosis Brain.” Acta Neuropathologica Communications 11(1): 69. doi:10.1186/s40478-023-01561-6.

Swanson, Molly E. V., Miran Mrkela, Clinton Turner, Maurice A. Curtis, Richard L. M. Faull, Adam K. Walker, and Emma L. Scotter. 2025. “Neuronal TDP-43 Aggregation Drives Changes in Microglial Morphology Prior to Immunophenotype in Amyotrophic Lateral Sclerosis.” Acta Neuropathologica Communications 13(1): 39. doi:10.1186/s40478-025-01941-0.

Tomasi, Giampaolo, Paul Edison, Alessandra Bertoldo, Federico Roncaroli, Poonam Singh, Alexander Gerhard, Claudio Cobelli, David J. Brooks, and Federico E. Turkheimer. 2008. “Novel Reference Region Model Reveals Increased Microglial and Reduced Vascular Binding of^11^ C-(*R*)-PK11195 in Patients with Alzheimer’s Disease.” Journal of Nuclear Medicine 49(8): 1249–56. doi:10.2967/jnumed.108.050583.

Tong, Junchao, Belinda Williams, Pablo M. Rusjan, Romina Mizrahi, Jean-Jacques Lacapère, Tina McCluskey, Yoshiaki Furukawa, et al. 2020. “Concentration, Distribution, and Influence of Aging on the 18 kDa Translocator Protein in Human Brain: Implications for Brain Imaging Studies.” Journal of Cerebral Blood Flow & Metabolism 40(5): 1061–76. doi:10.1177/0271678X19858003.

Vokali, Efthymia, Elodie Chevalier, Nicolas Dreyfus, Dorian Charmey, Tania Melly, Jacqueline Kocher, Monisha Ratnam, et al. 2025. “Development of [18F]ACI-19626 as a First-in-Class Brain PET Tracer for Imaging TDP-43 Pathology.” Nature Communications 16(1): 9358. doi:10.1038/s41467-025-64540-6.

Wijesinghe, Sasvi S, James B Rowe, Hannah D Mason, Kieren S J Allinson, Reuben Thomas, Davi S Vontobel, Tim D Fryer, et al. 2025. “Post-Mortem Validation of *in Vivo* TSPO PET as a Microglial Biomarker.” Brain 148(6): 1904–10. doi:10.1093/brain/awaf078.

Woollacott, Ione O. C., Christina E. Toomey, Catherine Strand, Robert Courtney, Bridget C. Benson, Jonathan D. Rohrer, and Tammaryn Lashley. 2020. “Microglial Burden, Activation and Dystrophy Patterns in Frontotemporal Lobar Degeneration.” Journal of Neuroinflammation 17(1): 234. doi:10.1186/s12974-020-01907-0.

Yaqub, Maqsood, Bart Nm Van Berckel, Alie Schuitemaker, Rainer Hinz, Federico E Turkheimer, Giampaolo Tomasi, Adriaan A Lammertsma, and Ronald Boellaard. 2012. “Optimization of Supervised Cluster Analysis for Extracting Reference Tissue Input Curves in (R)-[11C]PK11195 Brain PET Studies.” Journal of Cerebral Blood Flow & Metabolism 32(8): 1600–1608. doi:10.1038/jcbfm.2012.59.

